# Neural Correlates of Effects of Internally versus Externally Guided Partnered Rehabilitative Tango for People with Parkinson’s Disease

**DOI:** 10.1101/2021.07.16.21260219

**Authors:** Amrit Kashyap, Bruce Crosson, Venkatagiri Krishnamurthy, Keith M. McGregor, Ariyana Bozzorg, Kaundinya Gopinath, Lisa C. Krishnamurthy, Steve L Wolf, Ariel R Hart, Bing Ji, Marian Evatt, Daniel M. Corcos, Madeleine E. Hackney

**Author notes:** Corresponding Author: Madeleine E. Hackney, PhD, Associate Professor, Department of Medicine, Division of Geriatrics and Gerontology, Research Health Scientist, Rehab R&D Center, Atlanta VA Health Care System, 1841 Clifton Rd. NE, Atlanta GA 30329.

## Abstract

Restoring function to damaged neural pathways, or promoting compensatory strategies to overcome dysfunctional neural pathways have been topics of inquiry within motor rehabilitation. This study considers these topics in Parkinson’s disease (PD), where disruption within the striatal-thalamic-cortical (STC) circuits can cause impairment in internally guided (IG) movements. A related, but separate externally guided (EG) movement network, recruits the intact cerebellar-thalamic-cortical (CTC) loop to facilitate movement in response to sensory cues, and is effective in remediating motor function. Partnered dance with leading and following roles may be used as proxies for training IG and EG strategies respectively, in PD and can test which strategy is more effective in remediating effects of PD. Leaders determine variables associated with IG and communicate step amplitude, timing, and direction to the follower. Followers use an EG strategy to sense and interpret directional pressure cues from the leader and then enact an appropriate movement response. This study examines how IG and EG training strategy affects STC and CTC circuits and their behavioral outcomes to examine whether compensatory or direct entrainment of neural pathways is more effective in PD. Fifty-eight participants were recruited with mild-moderate PD (stages 1-3) and randomly assigned to EG, IG or non-dance education control group and assessed before and after 12 weeks of biweekly interventions. Participants were assessed with standard cognitive and motor behavioral measures and lay in a Magnetic Resonance Imaging (MRI) scanner while they tapped their foot under two conditions: internal (tapping a learned rhythm: IT) and external (participant taps in response to an assistant’s felt tap on the participant’s hand: ET) guidance. The foot-tapping data collected with an accelerometer were evaluated by analyzing the frequency spectrum to calculate amplitude and timing of the foot taps. The functional (fMRI) data were pre-processed (AFNI), registered (MNI), and analyzed for changes in activation using a general linear model in SAS and AFNI. Postintervention, both the EG and the IG groups showed clinically significant changes on disease severity, but the EG group showed improvements on cognitive, motor, and mood variables. The EG group also outperformed the IG group in the in-scanner task performance measured by the foot accelerometer. Imaging data revealed a significant increase in the EG group in the primary motor cortex lower limb region and the parts of the cerebellar circuits, particularly right Cerebellar Lobule VIIIa. The control group showed an increase in activity in the putamen compared to the IG and EG groups that could be due to a different compensatory pathway. All findings were corroborated using a region of interest approach examining the same pathways according to an atlas. Our results indicate that the most effective strategy for the PD participants involved external cues that increased activity in the compensatory CTC pathway, the primary motor region, and significant improvements on almost all behavioral measurements.

## 1. INTRODUCTION

Neurorehabilitation using motor therapies has come into increasing focus as an alternate treatment to pharmacological and surgical methods for neurological disorders (McKay et al., 2016). These therapies can be implemented in parallel with traditional strategies as part of a comprehensive treatment plan (Dong et al., 2016). Successful motor therapies in Parkinson’s Disease (PD) have focused on improving motor symptoms by engaging participants in a wide range of motor activities (e.g., mobility training, dance, tandem biking, tai chi) that improve lower limb control, postural stability, motor planning and sensory motor integration (Morris et al., 2009; Amano et al., 2013). These programs use a mix of internally guided (IG) and externally guided (EG) movement strategies, both of which have evidence to support their application in rehabilitation. These strategies also may repeatedly engage crucial neural circuits that can be retrained and strengthened to offset the progression of PD. Either rehabilitative treatments may improve the striatal circuits that are damaged, or they target a compensatory parallel circuitry using the relatively undamaged cerebellum. (Cerasa et al., 2006; Elsinger et al., 2006; Yu et al., 2007; Hackney et al., 2015; Drucker et al., 2019). We consider these rehabilitative mechanisms in patients with PD. A better understanding of the mechanisms contributing to beneficial exercise, could improve the design of motor rehabilitation interventions for symptoms (e.g., freezing, bradykinesia) and the various disease stages of PD.

PD often leads to motor deficiencies, e.g., gait initiation, planning and selecting movement, and postural instability (Galvan et al., 2015; Tsai et al., 2009; Eckert et al., 2006; Low et al., 2002; Wu et al., 2011). Due to dysfunction of the striato-thalamo-cortical (STC) circuit, people with PD may have particular difficulty with IG tasks (Eckert et al., 2006; Low et al., 2002; Wu et al., 2011). These deficiencies may be overcome by *internally guided* (IG) strategies, which require an increased focused on preparing self-initiated movement by mentally rehearsing or motor planning (Morris et al., 2009). Reducing complex movements into simpler elements may facilitate motor performance. Employing a movement strategy with increased focus on movement plans, mentally rehearsing or preparing for self-initiated movement is helpful. For example, focusing on critical movement aspects (e.g., longer steps, quicker movements) helps individuals with PD to achieve nearly normal speed and amplitude (Rochester et al., 2006). Abundant evidence also supports *externally guided* (EG) movement strategies, which encourages the initiation of motor responses from sensory cues using cerebellar circuitry (Elsinger et al., 2006; Hackney et al., 2015). Benefits of visual and auditory cueing in behavioral studies have been demonstrated for people with PD (Rocha et al., 2014), and such cues are often incorporated in physical therapy, (O’Sullivan et al., 2014). Tactile cues have similar results, are processed even faster, with less attentional demand, and more efficiently than visual and auditory cueing (Ivkovic et al., 2016; Rabin et al., 2015; Rabin et al., 2010; van Wegen et al., 2006). Previous studies have shown that providing cues within motor therapies significantly improves measurable clinical variables important in PD such as stride length, and reducing timing errors (Rocha et al., 2014; Rabin et al., 2015; Ivkovic et al., 2016, Bella et al., 2015).

Most motor movements effective in improving PD are complex and involve both striatal and cerebellar circuits (Dong et al., 2016; Hackney et al., 2015). However, since the striatum is damaged in PD, designing an experiment to test if there is an effect in behavioral and neural responses due to EG or IG training becomes important. Individually tested, IG training focusing on critical movement aspects (e.g., longer steps, quicker movements) helps PD participants recover nearly normal speed and amplitude when compared to age matched controls (Rochester et al., 2006). However, research has also shown that individuals with PD react to external cues faster than self-initiated IG movement (Ballanger et al., 2006). Comparative studies of IG and EG strategies are difficult to conduct, due to many differences in motor strategies that might act as potential confounders. To address this gap, we designed an experiment with a shared activity, partnered adapted Argentine Tango (Hackney et al., 2007; Hackney and Earhart 2010; McKee and Hackney 2013; McKay et al., 2016), that involves two distinct roles with defined goals and rules, the leader and the follower. Adapted tango dance uses a frame constructed by the embrace of arms between two dance partners, which serves an effective tactile conduit to pass information between each other during dancing. The leader self-initiates direction, timing and amplitude of movements and therefore engages in an IG strategy and may use striatal circuits more than cerebellar circuits. The follower’s movement is cued by receiving proprioceptive input from numerous sensory stimuli including visual, auditory and tactile cues from their partner. The follower task requires tactile and proprioceptive integration and may engage the cerebellar circuits more than the striatal circuits (Drucker et al., 2019). We aimed to control to the extent possible other potential variables e.g., number of sessions, the social environment, assistants, music, and dance steps. Although neither movement strategy exclusively activates one circuit, by comparing these two we create a gradient to see the effects of training the impaired circuit versus the compensatory circuit.

To evaluate the relative effectiveness of EG vs IG training for people with idiopathic, mild-moderate PD on striatal and cerebellar circuitry, we conduct a longitudinal randomized controlled trial for participants with PD by assigning them either as leaders or followers in adapted tango. A third, non-dance control group was recruited and engaged in partnered and interactive group learning and was compared to the dance groups. To measure participant neural responses before and after the intervention, we measured the brain activity of the participants using functional magnetic resonance imaging (fMRI) during a motor task. Two motor tasks were designed for the fMRI scanner, which involved foot tapping that was internally initiated with respect to a beat (IT) or externally initiated foot tapping conditions based on tactile cues provided from an assistant (ET). These two motor tasks have been used in our previous fMRI study to show that they selectively activate striatal and cerebellar circuitry and were used in motor assays in determining neural networks of the lower limb during EG and IG tasks in PD (Drucker et al., 2019; Dobkin et al., 2004). The participants were scanned before and after the intervention and were also assessed in a range of cognitive, motor, psychosocial, and PD clinical measures.

The purposes of the study wer to determine: the association between intervention (EG, IG and control) and changes in neural activation in various brain regions of interest (ROI) during task performance (ET, IT); the association between intervention (EG, IG and control) and the changes in the measured task (ET, IT) performance (timing, amplitude) as determined from the foot tapping data; and the association between neural changes, behavioral task changes and cognitive, motor, psychosocial, and PD specific function as a result of intervention. We hypothesized that leading and following train different neural pathways. Specifically, the following role may engage the CTC pathway more and lead to enhanced activation in that circuit; whilst the leader role may optimally engage the STC pathway but needs to use the CTC pathway in compensation.

## METHODS

The institutional review board at Emory University School of Medicine and the Research and Development Committee of the Atlanta VA Health Care System approved this work. Participants provided written informed consent before participating.

### 2.1 Participants and initial assessments

Fifty-eight participants were recruited, through multiple pathways: the Atlanta VA Center for Visual and Neurocognitive Rehabilitation (CVNR) registry, the VA Informatics and Computing Infrastructure database, the Michael J. Fox Foxfinder website, the Movement Disorders unit of Emory University, PD organizations newsletters, support groups and educational events and through word of mouth. The following selection criteria was used to screen participants. All PD participants were clinically diagnosed with PD by a movement disorders specialist. This diagnosis was based on the United Kingdom PD Society Brain Bank diagnostic criteria (Hughes et al., 1992). Participants were aged 40 and older and could walk 3 meters or more with or without assistance. No participants had contraindications to undergo a functional magnetic resonance imaging (fMRI) scan. Participants could hear above the pure-tone threshold (>40dB). Participants were excluded if they scored <18 on the Montreal Cognitive Assessment (MoCA) (Nasreddine et al., 2005; Hoops et al., 2009). Exclusion criteria also included peripheral neuropathy, untreated major depression, history of stroke, or traumatic brain injury. The Beck Depression Inventory-II (BDI-II) assessed depression and a score of ≥30, indicating severe depression, was a cutoff for the BDI-II. (Schrag et al., 2007). All participants were right-handed as verified by the Edinburgh handedness survey. Patients had unilateral onset of symptoms, displayed clear symptomatic benefit from antiparkinsonian medications, e.g., levodopa, and were in Hoehn and Yahr stages I-III (Hoehn and Yahr 1967). Patients who had a tremor score greater than 1 on the Movement Disorders Society Unified Parkinson Disease Rating scale (MDS-UPDRS) part III in either lower limb and/or moderate-severe head tremor were excluded. PD participants were tested in the OFF state, i.e. at more than 12 hours after their last dose of anti-parkinsonian medication.

Participants were randomly assigned to one of three groups with a computer-generated pattern (randomizer.org). Participants attended twenty, 90-minute adapted tango classes as a leader or a follower, or twenty 90-minute health education classes, within a 12-week period. Participant characteristics at Baseline are summarized in Table 1. Trained raters administered measures according to standard procedures. Table 1 shows clinical and demographic characteristics and baseline measure of cognitive, motor, disease, and psychosocial function of participants who completed 20 lessons. One-way ANOVA showed groups were not significantly different on any of the baseline characteristics

**Table 1:** Participant Demographics and Clinical Characteristics. Participant Characteristics. Values are presented as mean and standard deviations or in counts. MDS-UPDRS = Movement Disorders Society Unified Parkinson’s Disease Rating Scale. One-way ANOVA showed groups were not significantly different on any of the characteristics, p>.05

| **Table 1: Participant Demographics and Clinical Characteristics** |  |  |  |
| --- | --- | --- | --- |
|  | **IG (N=21)**<br>**(Leaders)** | **EG (N=20)**<br>**(Followers)** | **Control**<br>**(N=17)** |
| **Demographics** |  |  |  |
| **Age (years)** | 69.8(5.8) | 67.6(8.9) | 66.6(10.0) |
| **Sex (M/F)** | 14/6 | 12/9 | 12/5 |
| **Education (years)** | 16.4(2.8) | 17.1 (2.3) | 16.4(1.8) |
| **Duration with PD (years)** | 6.2 (5.0) | 6.1 (5.0) | 6.4(4.0) |
| **Composite Physical Function (/24)** | 20.0(4.5) | 18.8(6.0) | 18.4(4.8) |
| **Assistance needed to accomplish**<br>**Activities of Daily living (v/n)** | 2/18 | 6/15 | 2/15 |
| **Number of Comorbidities** | 3.6 (1.5) | 2.9 (1.8) | 3.9(1.9) |
| **Number of Prescription Medications** | 6.2(3.8) | 4.4(2.7) | 6.1(4.2) |
| **Activities Balance Confidence Scale** | 76 (17) | 76 (21) | 73 (27) |
| **Physical Activity Scale for the Elderly** | 114 (68) | 100 (54) | 93 (87) |
| **Montreal cognitive assessment (/30)** | 25.2(3.8) | 25.5(3.4) | 26.7(2.8) |
| **Beck Depression Index (/64)** | 10.7(5.0) | 11.5(8.4) | 13.5(8.9) |
| **PD Variables** |  |  |  |
| **MDS-UPDRS I** | 13.0 (5.4) | 11.5(7.2) | 12.4(7.3) |
| **MDS-UPDRS II** | 13.6(7.4) | 13.1(8.4) | 15.9(7.3) |
| **MDS-UPDRS III** | 33.8(10.6) | 35.1(11.3) | 35.7(11.4) |
| **MDS-UPDRS IV** | 3.1(3.5) | 3.0(3.1) | 3.7(3.9) |
| **Freezing of Gait** | 5.0(4.0) | 7.3(6.1) | 7.2 (5.2) |
| **Hoehn & Yahr Stages** | 2.1(.6) | 2.2(.7) | 2.2(.5) |
| **Table 1.** Participant Characteristics. Values are presented as mean and standard deviations or in counts. MDS-UPDRS = Movement Disorders Society Unified Parkinson's Disease Rating Scale. One-way ANOVA showed groups were not significantly different on any of the characteristics, $p>.05$ | | | |

### 2.2 Intervention Adapted Tango and Educational Control

Adapted tango is an adapted form of Argentine tango dancing designed to target the movement impairments of individuals with PD through modification of the tango frame and steps. Composed of simple steps, tango involves frequent movement initiation and cessation, multi-directional perturbations and varied rhythms. Participants are instructed to focus on trunk control and stepping strategies, coordination, kinesthesia, their partner, their trajectory, and aesthetics of the steps. More details about adapted tango format are provided (Hackney and Earhart 2010; Hackney and McKee 2014). Adapted tango methods are also delineated in an adapted tango manual, which describes aging-specific motor impairments, fall risk and prevention, partnering enhancement, and rhythmic entrainment, and a 20-class syllabus. The syllabus in the manual has is effective for teaching PD participants (McKee and Hackney 2013).

Class sizes were limited to 10 pairs of participants with PD and healthy partners to maximize safety. PD participants danced with healthy partners only and switched partners every 15-20 minutes, widely practiced method intended to enhance learning in partnered dance lessons. During classes, participants danced only their assigned roles: either IG (leading) or EG (following). Undergraduate and graduate students participated in all interventions as partners to the PD participants. Participants in IG training danced the leader role and determined steps as well as the direction, timing and amplitude of each successive step. Participants in EG training danced the follower role and attended to tactile pressure cues on their elbows and forearms from their partner’s hands, palms and forearms for movement direction, timing and amplitude of steps. Both IG and EG participants heard the same music at the same time, which controlled for the influence of auditory cues.

Participants in the education group engaged in health education lectures for 1 hour, and then participated in 1/2 hour of interactive small group and partnered discussion. These sessions employed learning techniques for memory retention. Discussion was strongly encouraged through the lecture. Medical students as well as faculty and experts from Emory School of Medicine and other local universities or organizations presented information on latest research in health topics relevant to an older adult population. Student volunteers participated in these sessions by moderating the lectures, assisting with the presentation of information and leading discussions. Participants in this training were instructed not to change their habitual exercise routines. More details about the education group can be found here: Dillard et al., 2018.

### 2.3 Behavioral Measurements

The following measures were administered at both preintervention (baseline) and postintervention timepoints. PD participants were administered the Movement Disorders Society UPDRS (MDS-UPDRS) parts I-IV (Goetz et al., 2008). Participants were evaluated over a range of motor and cognitive behavioral tasks. They completed psychosocial questionnaires. Specifically, participants were administered the Six Minute Walk Test, the Fullerton Advanced Balance and the Brooks Spatial Memory, and Corsi Blocks test.

The three groups were well matched in PD characteristics, age, sex, education, activity level, physical function levels/ability to perform ADLs, health metrics related to falls (number of comorbidities, number of prescription medications), cognitive function, balance confidence, and depression levels (Table 1.)

### 2.4 Foot Tapping assessment during Imaging

The task was performed per methods described by Drucker et al., 2019. Laying supine, participants performed a foot-tapping task with an orthopedic wedge under the knees to maintain a 90º angle and isolate ankle movement (Figure 1A). They wore a custom-made instrumented ankle orthotic on the leg more affected by PD based on leg agility and foot tapping items of the MDS-UPDRS-III. For equal bilateral rating, patient-reported side of parkinsonian symptom onset was factored. Participants were told to dorsiflex, then plantarflex till they reached a stop. The arc between stops, the largest possible range of motion (ROM) was 20 degrees. The orthotic affixed to the lower extremity incorporated position measurement, and sampled foot position continuously (2000 Hz). Custom software (BioMaq) built with LabView (National Instruments, Austin, TX) acquired data regarding timing and amplitude of foot tapping. Before scanning, participants learned a 4 second rhythmic sequence designed to approximate a basic rhythm of the Argentine tango dance: slow, slow, quick, quick, slow (Figure 1B). The tango rhythm was chosen to present a more complex rhythm than previous studies and for its current and future use in rehabilitation, e.g., with adapted tango therapies (Hackney et al., 2015). Intervals between taps were 1 s, 1 s, .5 s, .5 s, 1 s (Figure 1C). All participants were trained approximately 20 minutes on the task while lying on the scanner table.

**Figure 1.**
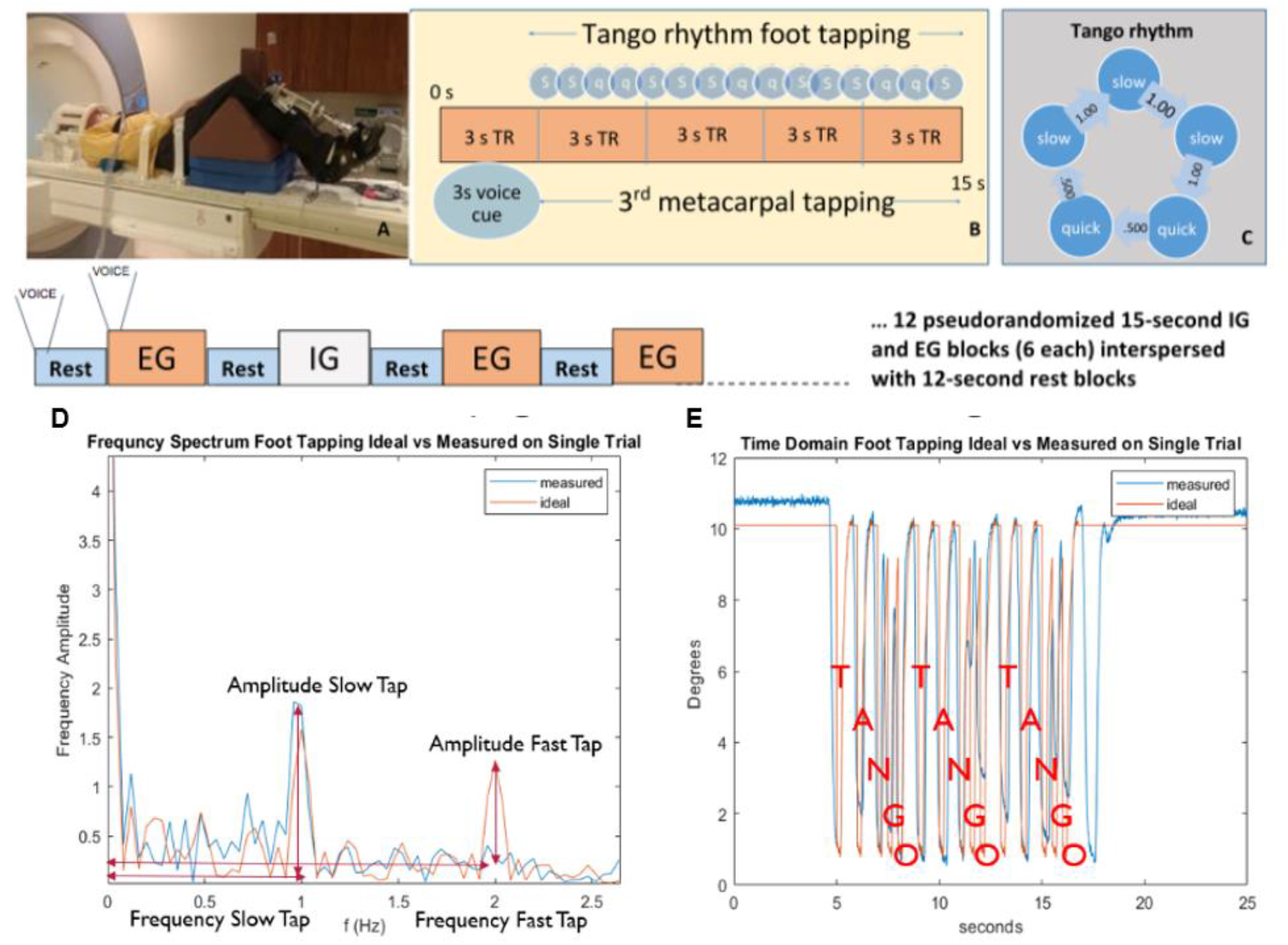
Foot Tapping Motor Assessment. Activation Paradigm and Participant MRI Set Up: Schema of the Foot Tapping Test. A participant prepared for the task lies in the scanner (A). Foot tapping sequence within each active block (B). During task blocks, the participant tapped their foot to the Tango rhythm: Slow, Slow, Quick, Quick, Slow. (C). The timing of each tap to a single tango beat is shown in C, with the rhythm starting at the top of the pentagon and going clockwise. A Fourier transform of the entire block of foot tapping is shown in (D), where red represents the ideal from the time domain, and blue is an observed trace. The ideal time series of 3 consecutive tango beats representing a single task block is shown in red (E), whereas the blue trace represents an observed response. Voice= voice cue.

Examiners verified that the participant fully understood the task and could perform the ET (externally timed), IT (internally timed) and Rest conditions proficiently and as instructed before beginning the scanning session. **ET condition blocks** : participants tapped their foot immediately after detecting a tap on their 3rd metacarpal head from an examiner listening to an audible version of the rhythm via headset. The examiner was placed just outside the scanner bore next to the patient table on the affected side of the participant. The examiner ensured access to the participant’s hand prior to the scan. This examiner was extensively trained to synchronize their hand taps to the auditory timing provided with the headset. **IT condition blocks:** Participants tapped the rhythm as practiced before the scan without receiving any prompting tactile cues on their hand. The examiner instead tapped the 3rd metacarpal head *after* each participant foot-tap to control for the experience of tactile sensation in the ET and Rest conditions. Participants were told to not pay attention to the taps on their hand during the IT condition. **Rest condition :** Examiners tapped the participants’ 3rd metacarpal head at 1 Hz to control for the experience of the tactile sensation during the IT and ET conditions. There were twenty-four 15s blocks of ET, and twenty-four 15s blocks of IT tapping across four runs. Block order was randomized in each functional run. Throughout the first three seconds of each block, a voice cue indicated whether the participant would experience an ET or IT trial. With 12 second rest blocks interposed between task, tapping lasted almost 25 minutes (Fig. 1). Sequence order was counterbalanced across participants. A sound check was performed before scanning. Examiners contacted participants through the microphone during the scan if participants were not compliant with the task. The voice cue was provided because we wanted participants to keep their eyes closed throughout the scan so they would focus their attention on the sensation of the tactile cues. The 3 second voice cue of the task instructions were included as a regressor of non-interest in the model. Notes were taken by an observer in the scanning console room, regarding the task performance of participants.

### 2.5 Foot Tapping Accelerometer Processing

To denoise, foot tapping performance timesereis were low pass filtered at 20 Hz. The individual foot tapping aligned with the EPI sequence that was sampled at a 3 second interval. Sample foot tapping from the accelerometer after denoising is shown in a time series (Figure 1E). The corresponding T-A-N-G-O (slow, slow, quick, quick, slow) beat is annotated on the figure, where each of the peaks in deflection represent a foot tap. There are 3 trials in each task block resulting in fifteen foot taps within each task block (Figure 1E is one task block). To evaluate each task, a time segment centered around each of the 24 trials was taken and transformed into the frequency spectrum. The peaks around 1 and 2 Hz represent the fast and the slow taps respectively during the TANGO beat (Figure 1D). The amplitude and the peaks that were closest to 1 and 2 Hz were taken as behavioral measures of in-scanner task performance. The variables were evaluated in the frequency spectrum because of challenges with aligning the foot tapping data that was manually started at the beginning of e-prime and was not robust to minor changes in timing. Of the variables that were used to categorize the foot tapping task, the slow amplitude frequency is the most robust measure to noise.

Data from 5 IG participants and 1 Control participant were excluded from the foot tapping analysis, because data collection resulted in blank runs due to failure from the accelerometer. Some trials resulted in missing taps, where the participants did not tap their feet for undetermined reasons or bradykinesia led to non-detectable taps. These trials and blocks were marked to be censored for volumes for the fMRI statistical analysis.

### 2.6 Image Acquisition

The neuroimaging data were collected in the Center for Systems Imaging at Emory University on a research-dedicated 3T Siemens Trio scanner using a Siemens 12-channel head coil, both for functional and anatomical runs. In functional runs, 114 time slices of T2*-weighted echoplanar image volumes measuring BOLD contrast were collected using parallel imaging with an iPAT acceleration factor of 2. Scan sequence parameters were: 55 contiguous 3 mm slices in the axial plane, interleaved slice acquisition, repetition time (TR) = 3000 ms, echo time (TE) = 24 ms, flip angle = 90 degrees, bandwidth = 2632 Hz/pixel, field of view (FOV) = 230 mm, matrix = 76 x 76, voxel size = 3.0 x 3.0 x 3.0 mm. At the beginning of the run, the scanner acquired 3 TRs which were discarded automatically. An anatomical image was collected using a high resolution MPRAGE scan sequence with 176 contiguous slices in the sagittal plane, single-shot acquisition, TR = 2300 ms, TE = 2.89 ms, flip angle = 8 degrees, FOV = 256 mm, matrix = 256×256, bandwidth = 140 Hz/pixel, voxel size = 1.0 x 1.0 x 1.0 mm.

Care was taken to ensure that the whole of the cerebellum was covered by the bounding box of the fMRI data acquisition for all subjects. The MR imaging parameters of the EPI sequence employed during fMRI contained 55 axial slices with 3mm slice thickness. Coverage afforded by these parameters were enough to obtain whole brain fMRI data from all subjects.

### 2.7 Image preprocessing

Image preprocessing and statistical analyses were conducted with Analysis of Functional Neural Images (AFNI) and FSL 5.0 software. The scans were preprocessed through updated AFNI commands (Cox 1996; Jenkinson et al., 2012).

Slice-time correction and motion correction was performed on the functional volumes using the AFNI function 3dvolreg. Artifacts due to head motion, circulatory activity, and other noise sources were removed using FIX. The T1 images were first bias corrected for the B0 distortions. The anatomical image was skull-stripped using optiBET, corrected for intensity bias introduced by magnetic field inhomogeneity, and then transformed to MNI space using the FNIRT procedure, producing a nonlinear transformation based on local spatial properties (Lutkenhoff et al., 2014; Andersson et al., 2007). FNIRT was employed using boundary-based registration. Functional data were aligned to the anatomical image and transformed using the nonlinear transformation and smoothed using an isotropic, 3D, 6mm full-width-half-maximum (FWHM) Gaussian kernel. Signal intensities in each volume were scaled with z-transformation excluding the first six volumes from calculation of the mean and standard deviation, avoiding pre-steady-state outliers.

In addition to the preceding standard steps, the brain images of those patients who were more affected on the left side and had to tap with their right foot were flipped hemispheres using methods previously published, so that the left motor cortex would be active for all participants (Schwingenschuh et al., 2011; Drucker et al., 2019). Because this population is clinical, TRs with head movement exceeding .5 mm were excluded. As stated above, TRs associated with the foot tapper in which the participant was noncompliant and did not move were also censored.

### 2.8 Statistical Analysis of Neural Data

To estimate the association between the intervention and the neural changes, a general linear model (GLM) model was used to estimate the group by time changes for the neural activation during task onset. The differences were estimated using two different approaches. The first approach models the neural activation using AFNI 3dDeconvolve GLM command for each individual before and after intervention separately and then estimating the group by time differences using the beta weights of the individuals and another GLM model performed using the AFNI MVM command (Chen et al., 2012; Cox 1996). The second approach was used to test ROIs related to the hypotheses. The signal was averaged according to the Brainnetome atlas at each ROI, and then using a GLM model tested the group by time effects of intervention on the ROIs using Statistical Analysis Systems (SAS) (Fan et al., 2016, SAS Institute. 2016). Unlike the AFNI GLM approach, which first estimates the betas and then the group effects, in the second, atlas approach the statistical power is marginally increased by running everything in one model. The primary drawback is that the AFNI approach is tested on a voxel by basis and is tested at a higher spatial resolution than using SAS.

Fifty-six patients were scanned both before and after 12 weeks of intervention; however, processing was not possible for one EG participant and two Education participants due to registration errors. Therefore fifty-three patients’ scans were included in neural analyses.

### 2.8 Voxel Analysis using AFNI

The AFNI software commands 3dDeconvolve + 3dREMLfit, were used to extract the beta weights for each regressor (Cox 1996). A detailed description of the GLM regressors is plotted using AFNI below in Figure 2 along with the annotation with respect to the alignment of the task. For each scan, four contrasts were modeled using tents, comprised of the two tasks (ET and IT) and two regressors of non-interest, the voice cue, and the timepoint after the task labeled as ‘transition’. To estimate a single beta, all regressors under the task bracket are summed together and represent the area under the curve during activation. The voice and transition label timepoints that are the most variable and most dependent on the alignment of the system. Modeling both as separate regressors improved accuracy in the model. In addition, three autocorrelation terms (linear, quadratic and cubic), were used to model each time series scan to mitigate effects of scanner drift (Cox 1996). Volumes in which participants did not move their feet during an ET or IT block as measured by the accelerometer were censored from the analyses. Four scans, with 12 task blocks in each scan, were used to estimate task activation beta values at each voxel, for each participant at pre and post intervention timepoints. These estimates of betas were then included into another GLM model, to estimate group by time values.

**Figure 2.**
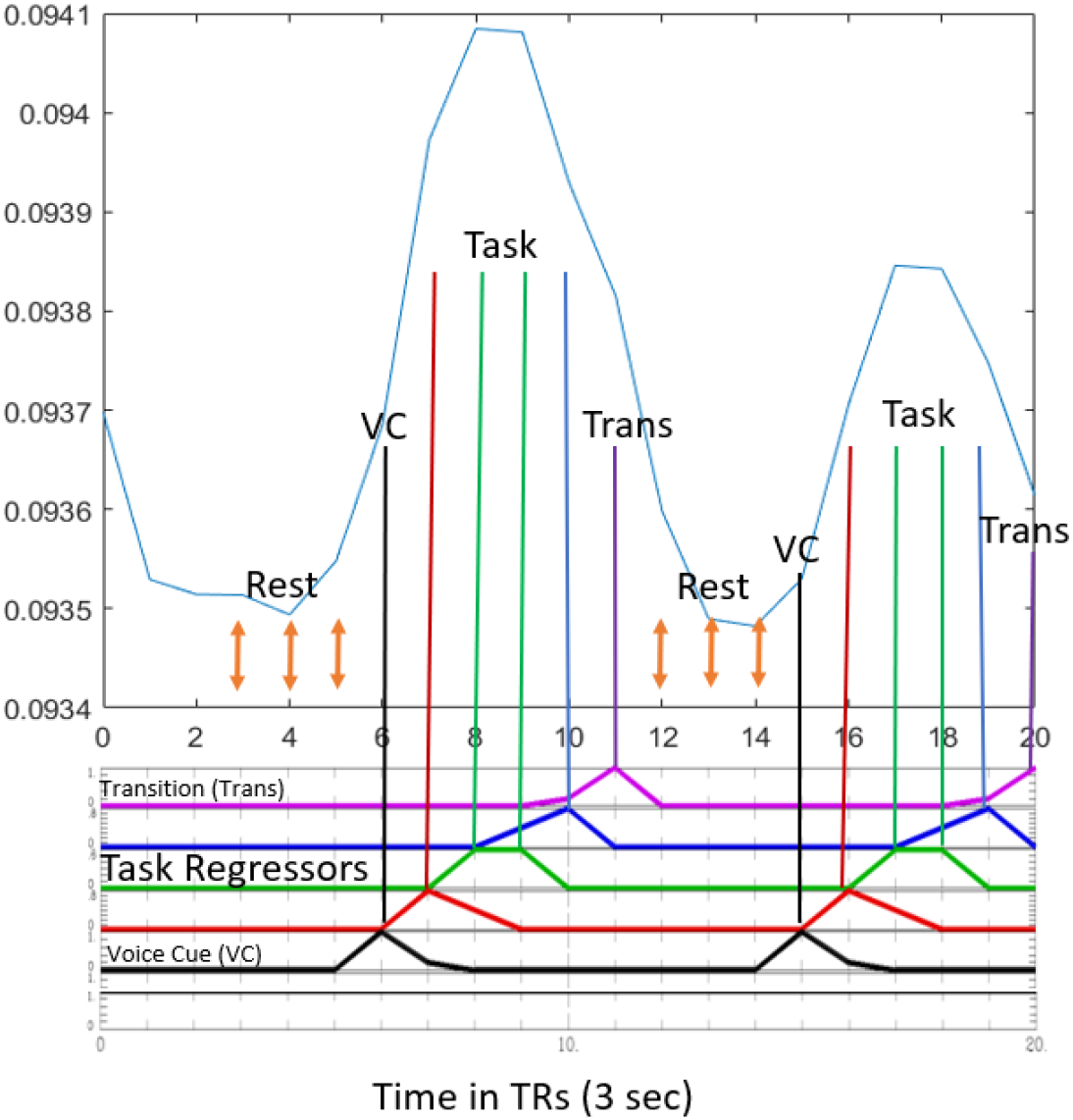
Average Time Series Response from Primary Motor Area of the Lower Limb. The average task response is plotted in the primary left lower limb motor cortex for the first 20 TRs averaged across all the subject scans prior to intervention. The alignment of the regressors for the general linear model is shown. The four TRs during task activation (12 sec) are estimated using three TENTs in AFNI and completed separately for the ET and IT tasks. The TR before and after the task are regressed out (shown as voice cue in black, and transition in purple). All unlabeled TRs constitute the baseline measurement. For SAS computation the tents are replaced with blocks. The first three TRs are not used in the computation.

For baseline analysis, the activation of the task networks due to the foot tapping for each ET and IT task was processed using 3dMEMA as suggested by AFNI software and output the group by time effects at a voxel wide level (Chen et al., 2012). To test for the changes due to intervention and by group, a more complex 3dMVM was used to compare the betas before and after activation due to differences in intervention group EG, IG, and Control. The 3dMEMA approach was not used here as it does not allow more nuanced models of statistical comparisons across multiple variables as 3dMVM (Chen et al., 2014). To control for false positives and set an appropriate cluster threshold, the residuals from the 3dREMLfit were fed into 3dFWHM -ShowMeClassicFWHM option for each individual scan and averaged together to generate one set of spatial autocorrelation values. To determine the significant threshold of these clusters, the 3dClustSim command with the -fwhmsxyz option and the spatial correlation was used to estimate the size of the clusters for a given uncorrected p value. This method is published in response to Eklund et al., 2016 and provides slightly more relaxed cutoffs (Gopinath et al., 2018). The Eklund et al., 2016 method yielded significant clusters during task activation but yielded no significant clusters for the much smaller changes due to motor intervention.

For the initial task activation estimates before intervention, the uncorrected *p* value was set to 0.0001 and the cluster significance was set to *q* = 0.01. These values gave a cluster threshold of 48 voxels for cortical regions, 21 voxels for cerebellar, and 8 voxels for striatal regions. At this threshold only the baseline task activation yielded significant differences between tasks. Group level changes due to intervention tested with 3dMVM yielded few significant clusters even when relaxed to p=0.005. As the cerebellum and the striatum have smaller grey matter regions and are the most important regions in the hypothesis, a mask was used to calculate the voxel thresholds for these regions (Poldrack 2007). The thresholds for the Cortex, Cerebellum and the Striatum are listed in Table 2. At voxel significance *p*= 0.005 and cluster significance *q*= 0.05, clusters that were larger than 164, 79, and 33 in the cortex, cerebellum and striatum respectively were considered significant for group by time differences.

**Table 2.** Group Cluster Thresholds. Group Cluster thresholds (number of voxels) calculated via the Gaussian distributions from the residuals on individual data and then averaged (Gopinath et al., 2018). The variable p represents the voxel wide significance, whereas the variable q represents the cluster thresholds. Values at p = 0.0001, q=0.01 were used to determine baseline activation rates. For the group by time comparison, the threshold was relaxed to p =0.005, and p = 0.05.

| | $q=0.05$ | $q=0.02$ | $q=0.01$ |
| --- | --- | --- | --- |
| **Full Brain** |  |  |  |
| $p = 0.005$ | 163.4 | 187.0 | 206.6 |
| p = 0.0001 | 35.5 | 42.7 | 48.1 |
| **Cerebellum** |  |  |  |
| p = 0.005 | 78.5 | 96.0 | 110.0 |
| p = 0.0001 | 11.1 | 16.5 | 21.2 |
| **Striatum** |  |  |  |
| p = 0.005 | 32.9 | 43.9 | 52.7 |
| p = 0.0001 | 2.1 | 4.9 | 7.6 |

### 2.9 ROI Approach

Since our hypothesis involves specific regions of interest (ROI) in the CTC and STC circuitry that are targeted by the intervention, an atlas brain parcellation was used to average and represent timeseries of interest in these regions (Brodman 1909; Fan et al., 2016) This approach improves the signal to noise-ratio of the timeseries by averaging regions; however at the cost of spatial resolution. The Brainnetome atlas was used to parcellate the brain activity into 274 regions of interest (ROIs) (Fan et al., 2016). The results display the ROIs that were determined by our a-priori hypothesis, but the analysis was run on all 274 brain regions. The atlas was first registered to the Montreal Neurological Institute (MNI) standard space. The atlas was down sampled from a 1mm resolution to the voxel resolution of our scans (3mm). After averaging the voxels, the ROI timeseries were then band passed filtered [0.01-0.1 Hz].

To estimate the group by time differences in these task ROI timeseries, a GLM mixed model implemented in SAS was applied on the timeseries data (SAS Institute, 2016). This model combines all data in one step and provides greater statistical power, compared to the AFNI approach. The pre and post time series of each of the participants were included in a single model to estimate the group by time changes due to each intervention. The GLM model uses the standard block-based task design where each time point is either labeled as ET when the external cue is present, IT when the internal cue is present, and the rest condition when they are both absent and the voice and transitions regressors (Figure 2). The only difference with the AFNI labeling in Figure 2 is that this process uses block activation instead of AFNI tents. The three intervention groups are encoded in the two variables EG and IG and the control group is represented when both variables are zero. The T variable corresponds to time and refers to timepoints either before or after the intervention. The rest of the variables were included to increase precision (Precision Terms), including three variables (t, t^2^, t^3^) dependent on time into each scan to account for temporal auto correlation and scanner drift (Cox 1996) To solve for the linear model, the SAS PROC MIXED command was used, which fits a different intercept beta for each subject, to account for individual variation and repeated measures. The GLM independent factors are shown in Equation 1 and all possible interaction terms are explicitly modeled as variables between the two tasks (ET, IT) and the three groups (EG, IG, control) and the precision terms. The regressors beta 6-7, represent the change in control groups for ET and IT activation, and combing them with beta 8-9 gives the change in activity for the dance groups. The final terms beta 10-14 represent all tertiary interaction terms. A sample test is shown below in Eq 2, to estimate the group by time effect of comparing EG to the IG group. To test the significance of the effect, the betas were compared against the two tailed t-distribution. We set our overall significance level at 0.05 percent. To correct for multiple comparisons, Bonferroni correction was used, which set the ROI threshold to 0.05/274 = ∼0.0001.

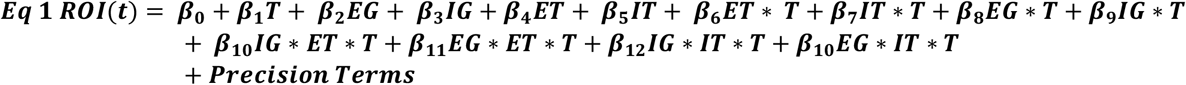

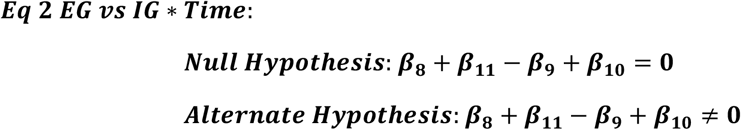

### 2.10 Participant Baseline Characteristics

Descriptive statistics were calculated for patient characteristics and behavioral outcome variables. Independent t tests were used to compare within group performance at pre and postintervention timepoints on the clinical variables. One way ANOVAs were used to compare baseline characteristics between groups.

### 2.11 Analysis of Foot Tapping Data

A GLM very similar to Equation 1, was used to test the changes in the neural data, as the foot tapping data were measured concurrently as the neural data and the design matrix for the foot tapping data analysis is akin to the design shown above. The variables (fast (quick tap) amplitude, slow (slow tap) amplitude, fast timing, slow timing) were averaged across the task block using the Fast Fourier transform (see Methods) rather than an ROI timeseries. Therefore, precision terms were not included with the timeseries variable, t. The model consisted of T (pre or post intervention) and the three groups (EG, IG, CN) and the two task conditions (ET, IT). The group by time effect for each task was then estimated for each variable (i.e, fast amplitude-the amplitude of the quick taps). The significance threshold was set at 0.05.

## 2. RESULTS

### 3.1 Behavioral Data

The mean and standard deviation of the behavioral variables pre/post intervention are shown in Table 3. Withing groups, all dance groups had significant changes in the MDS-UPDRS III score. The IG group improved on balance (Fullerton balance) and endurance (6MWT). The EG group improved on Freezing of gait, 6MWT, Corsi Blocks, Brooks Spatial Memory and Beck depression. The control group showed no significant improvement on any test.

**Table 3:**
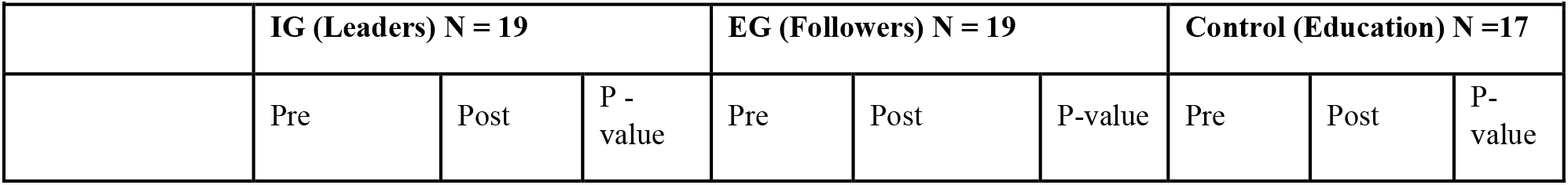
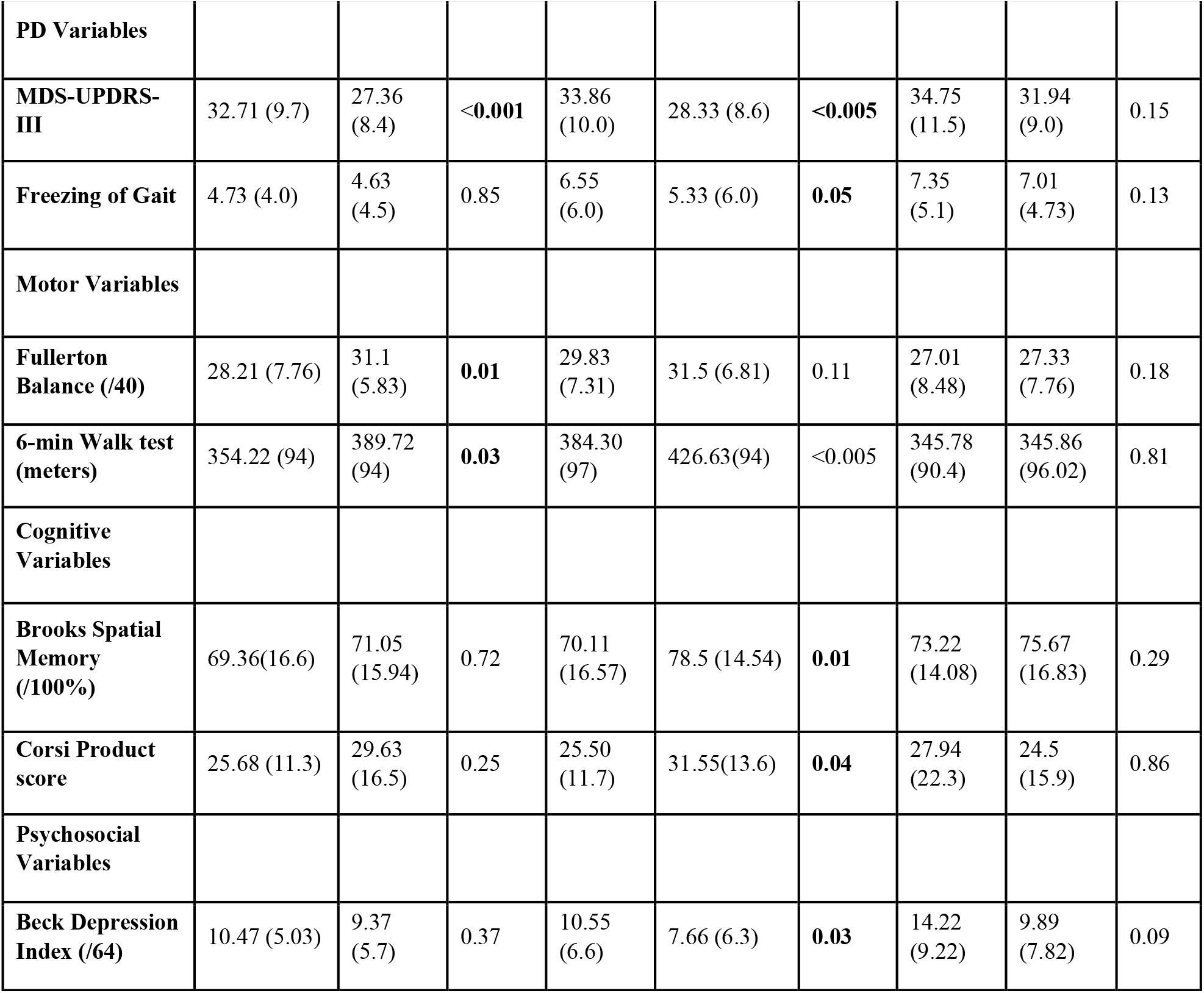
Pre and postintervention performance on clinical variables and within group significance of changes. Values are presented as means +/- standard deviation. The *p* values represent significance level of within group comparisons.

| Table 3: Pre and postintervention performance on clinical variables |  |  |  |  |  |  |  |  |  |
| --- | --- | --- | --- | --- | --- | --- | --- | --- | --- |
|  | IG (Leaders) N = 19 |  |  | EG (Followers) N = 19 |  |  | Control (Education) N =17 |  |  |
|  | Pre | Post | P - value | Pre | Post | P-value | Pre | Post | P-value |
| **PD Variables** |  |  |  |  |  |  |  |  |  |
| **MDS-UPDRS-III** | 32.71 (9.7) | 27.36 (8.4) | **<0.001** | 33.86 (10.0) | 28.33 (8.6) | **<0.005** | 34.75 (11.5) | 31.94 (9.0) | 0.15 |
| **Freezing of Gait** | 4.73 (4.0) | 4.63 (4.5) | 0.85 | 6.55 (6.0) | 5.33 (6.0) | **0.05** | 7.35 (5.1) | 7.01 (4.73) | 0.13 |
| **Motor Variables** |  |  |  |  |  |  |  |  |  |
| **Fullerton Balance (/40)** | 28.21 (7.76) | 31.1 (5.83) | **0.01** | 29.83 (7.31) | 31.5 (6.81) | 0.11 | 27.01 (8.48) | 27.33 (7.76) | 0.18 |
| **6-min Walk test (meters)** | 354.22 (94) | 389.72 (94) | **0.03** | 384.30 (97) | 426.63(94) | <0.005 | 345.78 (90.4) | 345.86 (96.02) | 0.81 |
| **Cognitive Variables** |  |  |  |  |  |  |  |  |  |
| **Brooks Spatial Memory (/100%)** | 69.36(16.6) | 71.05 (15.94) | 0.72 | 70.11 (16.57) | 78.5 (14.54) | **0.01** | 73.22 (14.08) | 75.67 (16.83) | 0.29 |
| **Corsi Product score** | 25.68 (11.3) | 29.63 (16.5) | 0.25 | 25.50 (11.7) | 31.55(13.6) | **0.04** | 27.94 (22.3) | 24.5 (15.9) | 0.86 |
| **Psychosocial Variables** |  |  |  |  |  |  |  |  |  |
| **Beck Depression Index (/64)** | 10.47 (5.03) | 9.37 (5.7) | 0.37 | 10.55 (6.6) | 7.66 (6.3) | **0.03** | 14.22 (9.22) | 9.89 (7.82) | 0.09 |

### 3.2 Neural Baseline Activation

The significant voxels of the task dependent neural activation for the ET and IT foot tapping task are plotted in Figure 3 and were determined according to methods section 2.8. The leftmost column for both tasks showed the largest cluster situated around the motor/ premotor area located on the medial motor cortex. During the task activation, the STC loop is also recruited as shown in the second column, with positive activation in the left putamen (44 voxels) and in the left thalamus region (46 voxels) for the IT task. The cluster in the thalamus is significant for both tasks, but the putamen is only significant in the IT task (Fig. 3, top row). The cerebellum is recruited for both IT and ET tasks, (middle column) and has bilateral activation in Lobules VI and V (top cluster in the cerebellum) and activity in the right lobules VIIIa/b (bottom cluster in the cerebellum). Activity in the left lobule VIIIa/b was significant for the IT task only. Bilateral activation occurred in the dorsal lateral prefrontal cortex (dlPFC), and in the pars triangularis as well which are known for motor planning areas as well. The time domain hemodynamic responses of these ROIs are plotted using the parcellations given by the atlas for the task onset in Figure 3 (A, B and C). The average time series response of the motor (M1 left lower limb), striatal (left putamen), and the right cerebellar (VIIIb) areas are plotted for both tasks with the time axis aligned with the task onset. The hemodynamic response function (HRF) is positive and significant (p < 0.0001) for all three areas during the motor task. There is no task difference (ET vs IT) in the HRF response difference in the putamen (Figure 3B); however, in the cerebellum VIIIb and M1 regions (Figure 3 A & C), the ET task shows an earlier and steeper response compared to the IT task.

**Figure 3.**
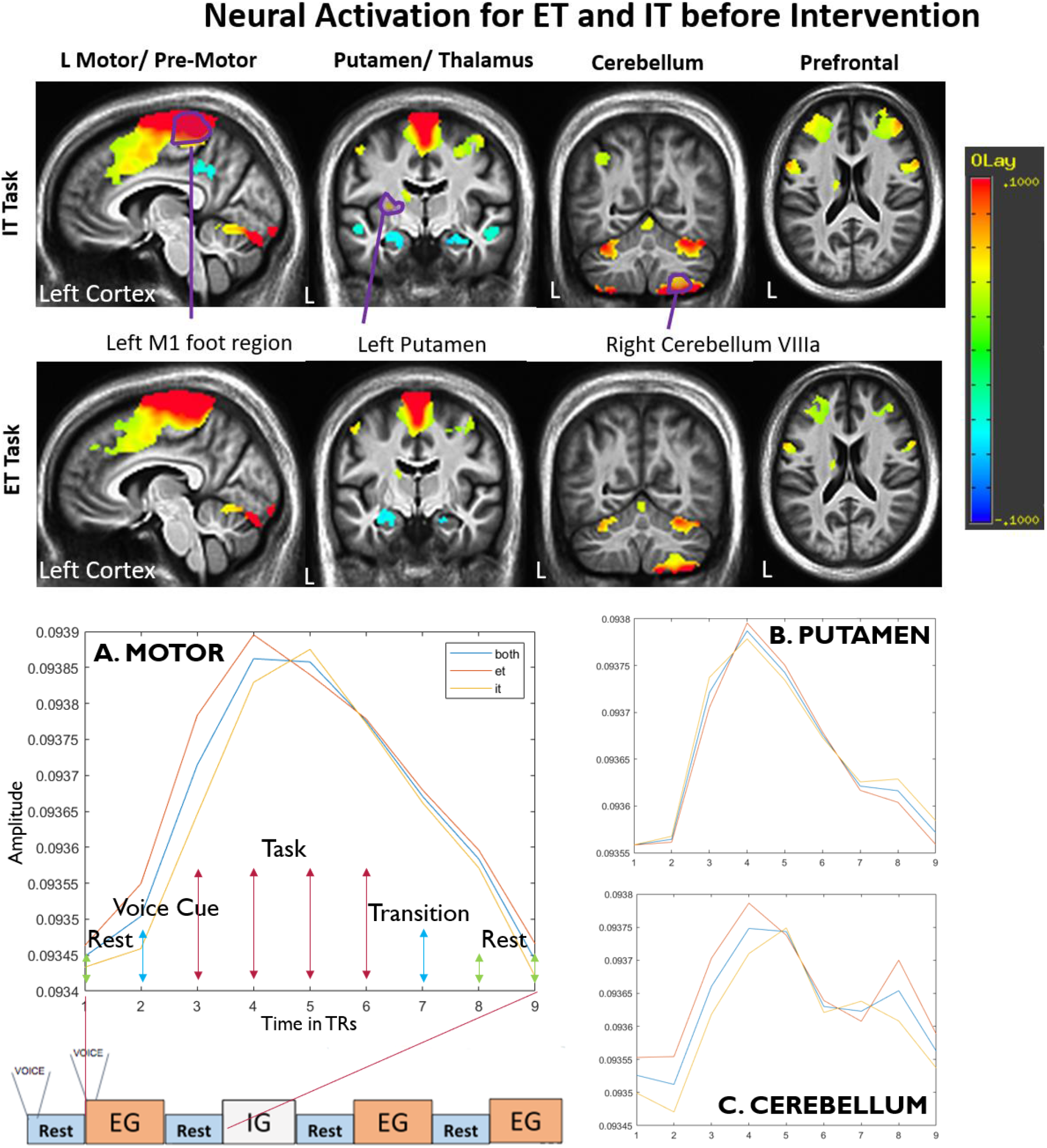
Neural Activation for External Tapping and Internal Tapping Preintervention for all participants: IT task (top row); ET task (bottom row). The leftmost column shows bilateral activation in the foot region of the motor/ premotor areas, bilateral deactivation in the posterior cingulate cortex, activation in the cerebellum and the visual system. The second column shows activation in the thalamus, putamen, M1 and SMA. The third column shows unilateral activation in the right cerebellar lobules VIIIa/b and bilateral in lobule VI and IV. Unilateral activation is also noted in the left precuneus for the IT task only. The rightmost, fourth column shows bilateral activation in the inferior frontal gyrus, left thalamus and left caudate. Bottom row: mean hemodynamic response function (HRF) in specific regions of the STC and CTC circuitry for both ET and IT tasks corresponding to the regions annotated above. Activation in left M1 aligns with the task activity, and slowly returns to baseline during rest. Larger and faster activation was noted in the right cerebellum (VIIIa/b) and the motor cortex during the ET task (red) compared with the IT task (yellow). Fewer differences were noted between HRFs in the left putamen.

### 3.3 Changes in Neural Activation Postintervention

The neural changes due to intervention involved several brain regions. As striatum and cerebellum are relatively smaller structures, the voxel/AFNI method with better spatial resolution was more sensitive to reveal smaller acute changes than the ROI method. However, in cortical regions such as the motor and supplementary motor areas where the changes were more widespread, the atlas approach performed better as small changes over large areas were averaged together. The full list of brain regions that changed after intervention is shown for voxels in Table 4 and for the ROI regions in supplementary Table 2. The following sections focus on changes in the STC, CTC and the motor areas are examined due to the intervention. The voxel results showed changes in the Cerebellum/Striatum while the ROI approach showed significant changes in the Cerebellum/Motor areas.

**Table 4.** Significant Voxel Clusters. Number of voxels that survived correction at p<0.005 are presented. Bolder values represent significant clusters q<0.05. Other areas of interest such as parts of either the striatal or cerebellar circuits are also included. CTRL=control.

| **Test** | **Brain Region** | **MNI** | **Voxels** | **Beta** |
| --- | --- | --- | --- | --- |
| **EG vs IG on ET** | Left Cerebellum Deep Cerebellar Nucleus | 12, 60, -40 | 26 | -1.27 |
| **EG vs IG on IT** | No changes |  |  |  |
| **EG vs CTRL on ET** | Anterior Cingulate Cortex | 4, -64, -8 | 75 | -1.39 |
|  | Right Cerebellum Deep Cerebellar Nucleus | -8, 60, -34 | 44 | -1.21 |
|  | Right Globus Pallidus | -18, 2, -6 | 29 | -1.6 |
|  | Left Cerebellum Deep Cerebellar Nucleus | 28, 70, -36 | 26 | -1.16 |
|  | Right Putamen | -30, 0, 2 | 20 | -0.9 |
| **EG vs CTRL on IT** | Left Precuneus | 12, 90, 42 | **194** | **-2.68** |
|  | Right Precuneus | -16,88,44 | 63 | -1.93 |
|  | Right Precuneus | -36,86,32 | 44 | -1.4 |
| **IG vs CTRL on ET** | Left Putamen | 22, -2, 10 | **75** | **-0.91** |
|  | Right Supplementary Motor Area | -6,-30,38 | 68 | -0.78 |
|  | Right Putamen | -30,-2,0 | **48** | **-0.97** |
| **IG vs CTRL on IT** | No changes |  |  |  |
| **IG pre vs post IT** | No changes |  |  |  |
| **IG pre vs post ET** | No changes |  |  |  |
| **EG pre vs post IT** | Right Dorsal lateral Prefrontal | -24,-54,30 | 99 | 0.76 |
|  | Pons Left | -10,20,-38 | **30** | **1.21** |
|  | Left Cerebellum Deep Cerebellar Nucleus | 16,58,-40 | 29 | -1.33 |
|  | Right Red Nucleus | -14,16,-24 | 26 | 1.52 |
| **EG pre vs post ET** | Left Cerebellum Deep Cerebellar Nucleus | 14,58,-40 | **91** | **-1.17** |
|  | Pons Right | 12,20,-40 | **68** | **1.73** |
|  | Sub Thalamic Nucleus | 4,18,-8 | 28 | 1.21 |
|  | Right Red Nucleus | -14,16,-24 | 26 | 1.52 |
| **CTRL pre vs post IT** | Right Putamen | -38,-4,-18 | **146** | **1.16** |
|  | Left Putamen | 20,-2,-8 | **70** | **0.71** |
|  | Subthalamic Nucleus | 10,10,-10 | **48** | **1.4** |
| **CTRL pre vs post ET** | Right Visual Cortex (V2) | -6,82,-12 | **200** | **2.11** |
|  | Pons nucleus | 2,38,-26 | **51** | **1.59** |
|  | Subthalamic Nucleus | 10,10,-8 | 24 | 1.1 |

### 3.4 Changes in the Striatum due to Intervention

The changes in the striatum after the intervention were most pronounced in the control group (Figure 4). The controls had increased activity in the putamen and large clusters were found to be significant in both left and right hemispheres (144 right, 70 left). Positive increases were also found in the internal globulus pallidus (Figure 4 top left and middle). These clusters partly overlap with changes in the temporal cortex especially on the right side, but excluding those regions there are still 70-90 voxels that localize to the putamen that significantly increased in the control group. In the dance groups, within group changes from pre vs post were not significant in the Putamen; however, these differences were significant compared with the control group (Figure 4 top middle and left). The ROI approach is not as sensitive in the smaller brain regions as the voxel approach and is not significant but showed a similar trend. The HRF functions (Figure 4 bottom row), indicate that the mean time trace has increased for the controls, no change for the EG group, and slightly decreased in the IG group.

**Figure 4.**
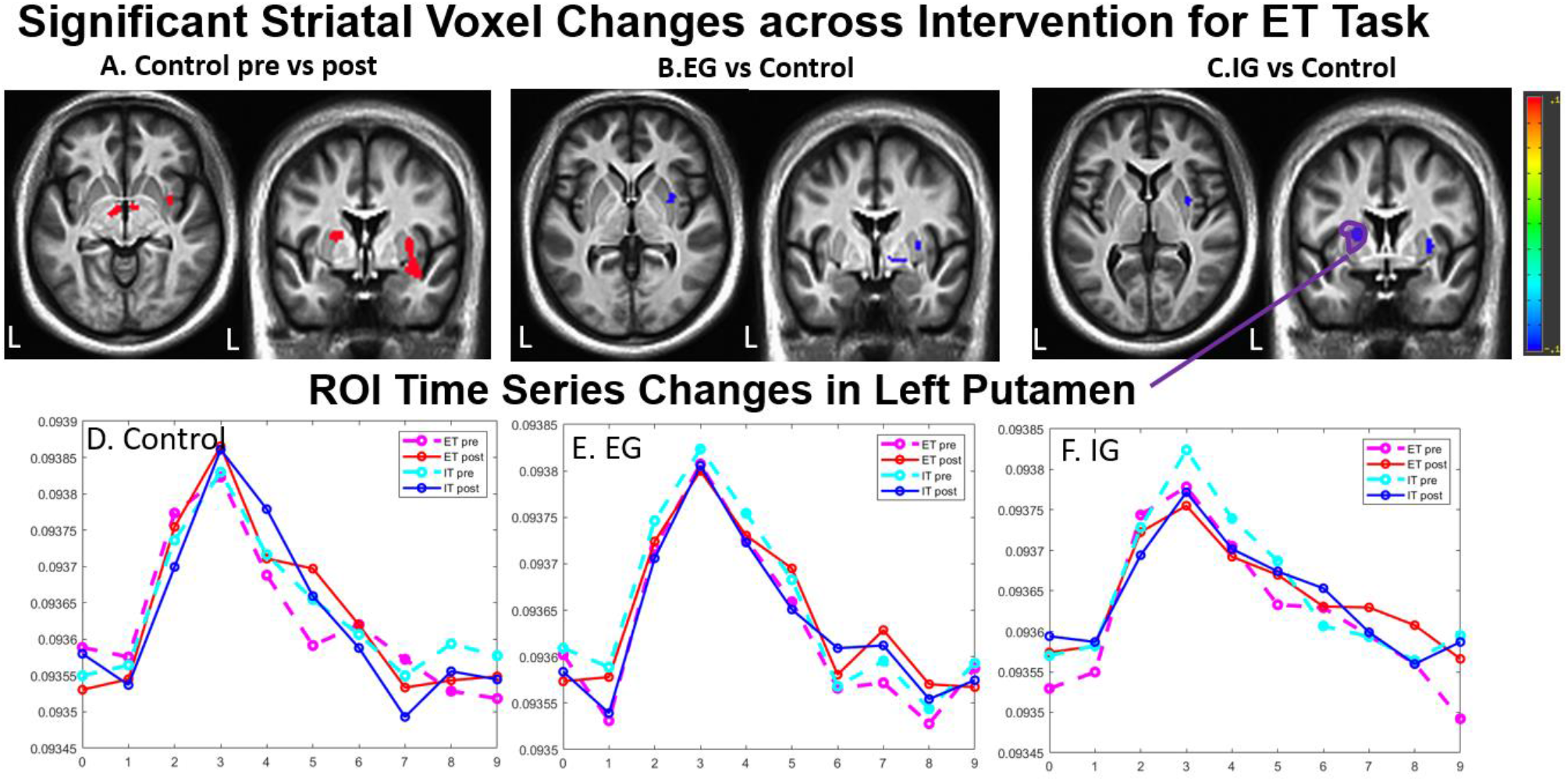
Pre vs Post changes in the Striatum: Controls had a significant bilateral increase in activity in the putamen and internal globus pallidus, after the three-month intervention, especially pronounced during the ET task (left panel). These changes in both the left and right Putamen are significant compared to dance groups (middle and right). ROI time traces (bottom row) in the left Putamen which was most active in the baseline task show the Control group increases activity, whereas the EG group remains unchanged, and the IG group shows a decrease of activity postintervention.

### 3.4 Changes in the Cerebellum due to Intervention

Changes in the CTC pathway were limited to the within group changes in the EG group, but the changes were significant in both voxel and atlas approaches. The ET task showed larger and significant clusters that survived thresholding (Figure 5 top row) than during the IT task, although the regions that change align across tasks (see Table 4). A large cluster of 73 voxels just outside of the cerebellum in the pons regions show a significant increase in activity in the EG group (Figure 5 middle two panels). The changes in the cerebellum showed decreased activity in the white matter areas, left Deep Cerebellar Nucleus (DCN) (cluster of 91 voxels in Figure 5 left two panels), to the EG intervention. The atlas ROI analysis corroborates these findings as it shows significant decrease in area VIIb Vermis, located in the near exact location (Figure 5 sagittal slice and Figure 6A). The areas in the Cerebellum that showed greatest change were contralateral to the side that was most active during baseline foot tapping but the cluster extends to both sides (Figure 3 Cerebellum). The timeseries of the area that was significantly active during baseline task, the right Cerebellum VIIIa, is plotted in bottom panels of Figure 5. It shows an increase in activity in the EG group while no discernable trend is noted in the other areas. The timeseries difference between groups is significant where the EG group shows more activity than the IG group in right Cerebellum VIIIa (Figure 6C).

**Figure 5.**
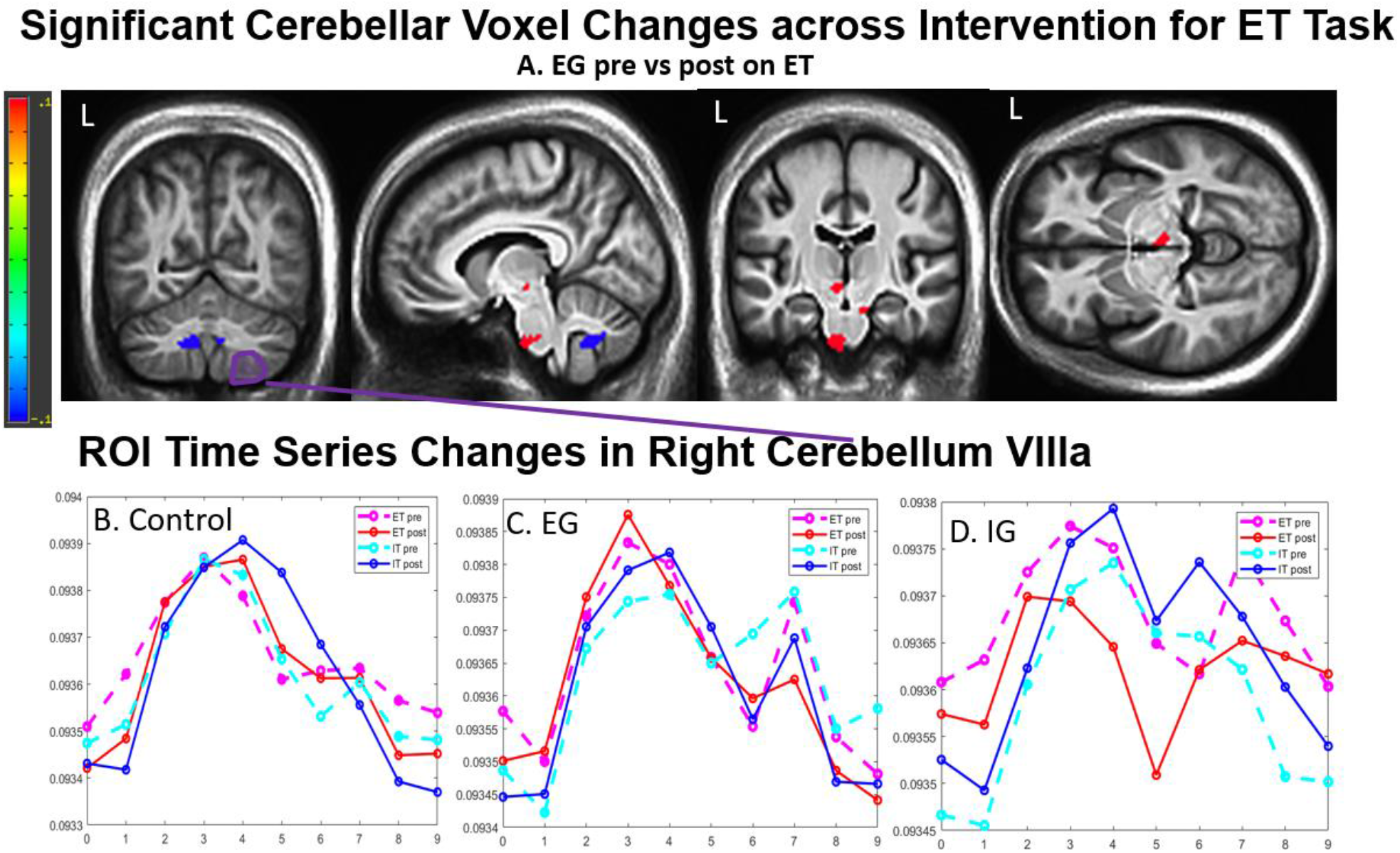
Pre vs Post changes in the Cerebellum: Within groups changes in the EG group from pre to post for both ET and IT task were noted, shown in a 91 voxel cluster that exceeded significance threshold located in the deep cerebellar area spanning the vermis (A: far left panel). Significant changes were also observed in the Pons (73 voxels). Two other areas in the motor pathway are shown to have non-significant clusters in the red nucleus (26 voxels) and left subthalamic nucleus (28 voxels). The atlas area closest to the white matter cluster, Cerebellum VIIb vermis shown in the same sagittal slice (Figure 6A) also shows a significant negative change in activity. The time traces show activity from the right Cerebellum VIIIa (see the same region in Figure 3) which is adjacent to the white matter clusters. The EG group has greater activity postintervention in both ET and IT HRF response. The control and IG group show no discernable trends. The EG vs IG ROI timeseries difference is also significant, where EG has greater post activity in right Cerebellum VIIIa (shown in Figure 6C).

**Figure 6.**
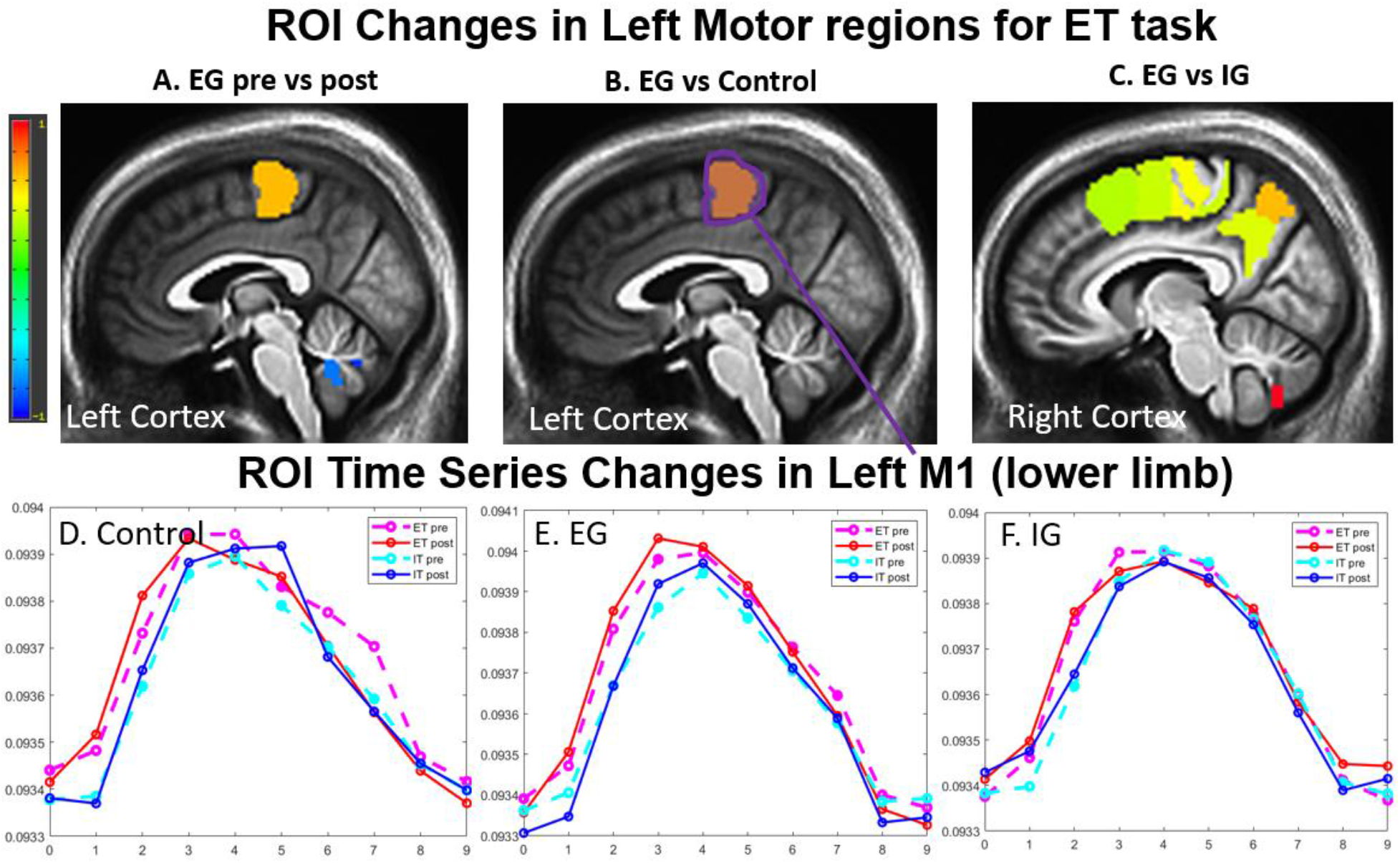
Pre vs Post Changes in Motor Cortical Areas: The atlas approach showed significant increases in the motor cortex especially in the atlas area that correspond to the left M1 lower limb area for the EG group. These differences are also significant when comparing between the EG group to IG and Control groups as well as within EG group (top). The changes were more pronounced in the ET task than in the IT task. The time traces from the M1 lower left limb are shown in Figure 6 D, E and F, where the EG unlike the other two groups show a faster and larger amplitude postintervention especially for the ET task.

### 3.5 Changes due to Intervention in the Motor Cortex

Significant changes in the motor cortex were only observed in the EG group using the ROI approach. These changes were significant when comparing within group EG changes and comparison to either the IG or the control group but were only significant for the ET task (Figure 6 top row). In all three comparisons, the left primary motor cortex in the lower limb region had increased in activity, which is contralateral to the right foot tapping task. The EG vs IG comparison showed the greatest number of changes also seen in the premotor areas and parietal areas. The M1 HRF functions showed larger and faster responses during the foot tapping task post intervention. The changes in these cortical regions were not captured in the voxel method.

### 3.6 Behavioral Foot Tapping Performance: Changes with Intervention

Performance during the in-scanner IT and ET foot tapping tasks at pre and postintervention is shown in the scatter plot in Figure 7 and summarized in Table 5. In the ET task, the EG group significantly improved in slow amplitude (amplitude of the slow taps) compared to Control and improved performance in fast frequency (timing of fast taps) compared to IG. The IG group decreased in fast frequency (timing of fast taps) of the ET task and slow amplitude (amplitude of slow taps) in the IT task compared to Control. In the IT task, the EG group improved in slow amplitude (amplitude of slow taps) compared to the IG group.

**Table 5.** Foot Tapping Variable Changes. Changes *p* < 0.01 are in bold. The values were calculated using the GLM model and represent the group by time difference between the two comparison groups (e.g., IG versus Control by Time). Performance at baseline approximated the ideal timing and amplitude, with room for improvement in all groups. These values show that postintervention, the EG group significantly improved compared to both the other groups, while the IG group decreased in performance.

| **Table 5. Foot Tapping Variable Changes** |  |  |  |  |  |  |  |
| --- | --- | --- | --- | --- | --- | --- | --- |
|  | Intercept | EG vs CTRL |  | IG vs CTRL |  | EG vs IG |  |
|  |  | val | pval | val | pval | val | pval |
| **Fast Amp IT** | 0.28 (0.01) | -0.03 (0.02) | 0.1141 | 0.029 (0.02) | 0.1687 | -0.06 (0.02) | 0.0240 |
| **Fast Freq IT** | 1.88 (0.02) | -0.04 (0.03) | 0.1325 | **-0.12 (0.03)** | 0.0001 | **0.073 (0.03)** | 0.0096 |
| **Slow Amp IT** | 0.72 (0.02) | **0.15 (0.04)** | 0.0006 | 0.08 (0.05) | 0.0651 | 0.06 (0.04) | 0.1484 |
| **Slow Freq IT** | 0.976 (0.01) | -0.015 (0.01) | 0.1284 | -0.015 (0.01) | 0.1560 | -0.00025 (0.01) | 0.9802 |
| **Fast Amp ET** | 0.34 (0.01) | -0.047 (0.02) | 0.0383 | -0.065 (0.02) | 0.0810 | 0.017 (0.02) | 0.4486 |
| **Fast Freq ET** | 1.91 (0.01) | -0.053 (0.02) | 0.0253 | -0.047 (0.03) | 0.0643 | -0.006 (0.02) | 0.8015 |
| **Slow Amp ET** | 0.89 (0.03) | 0.04 (0.06) | 0.4700 | **-0.2 (0.06)** | 0.0011 | **0.16 (0.06)** | 0.0059 |
| **Slow Freq ET** | 0.99 (0.004) | 0.01 (0.01) | 0.2300 | 0.017 (0.01) | 0.0741 | -0.0065 (0.01) | 0.4700 |

**Figure 7:**
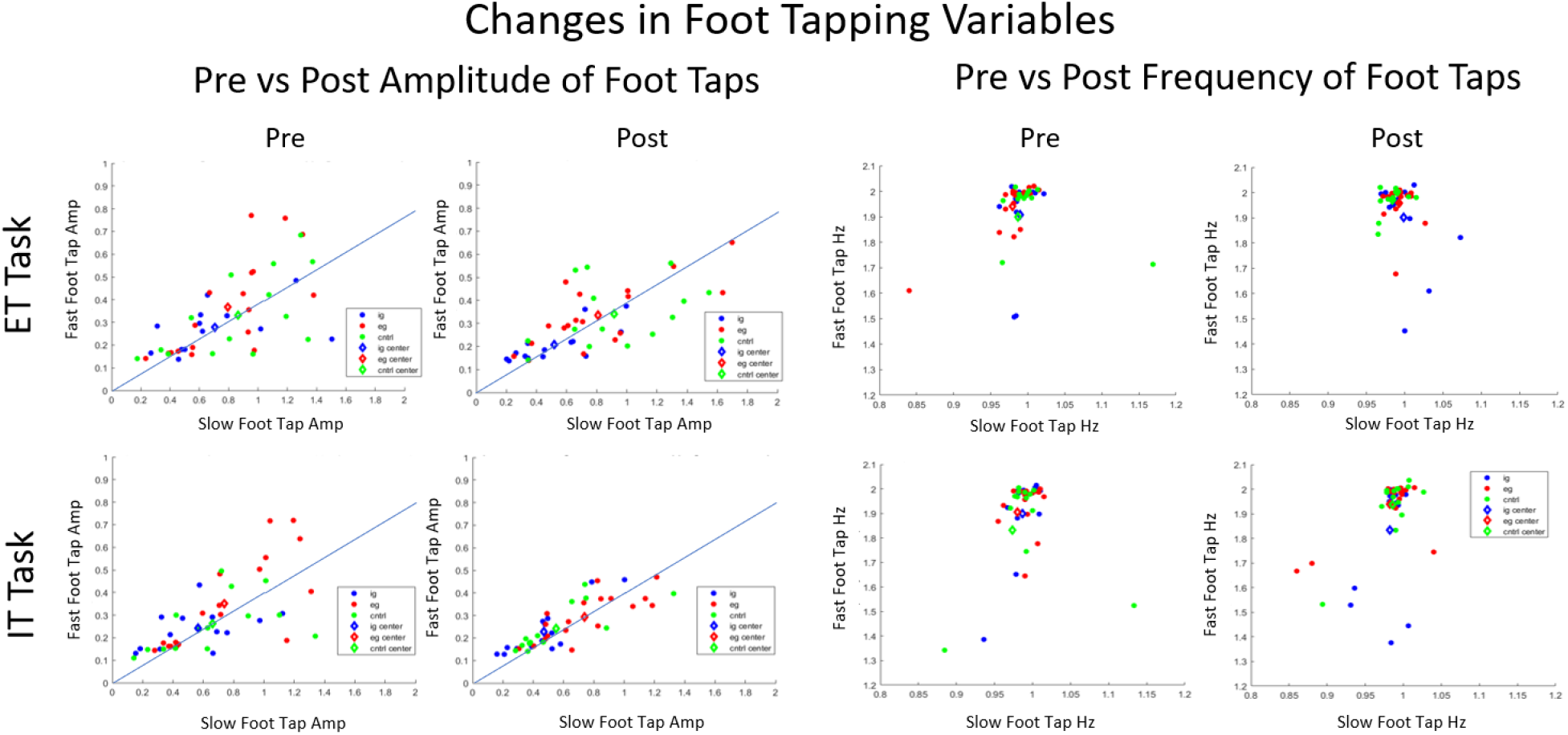
Changes in Foot Tapping Amplitude and Frequency Postintervention. Data presented are scatterplots of pre intervention (left) and post intervention (right) for Amplitude (A) and Frequency (B). Amplitude performance increases with the x and y axes. Ideal performance is a 2:5 amplitude ratio (plotted on the graph) for fast to slow tap amplitude. Frequency performance is best when clustered around x= 1 Hz (for slow taps), y= 2 Hz (for fast taps). Each dot in these plots represents the average of all 48 trials for one individual and the average of all groups is shown with a diamond. Preintervention, the amplitudes don’t have a relationship with the 2:5 ratio (r-squared 0.17, 0.04,0.27 for IG, EG, CTRL for ET, r-squared 0,24,0.20,0.17 for IG, EG, CTRL for IT). Post intervention the EG and IG groups have large r-squared with the line 0.61, 0.59 respectively for ET task and 0.53, 0.63, for the IT task. Post intervention the controls have low r-squared for the ET task 0.03 post intervention but for the IT task they show similar trends to the dance groups and have an r-squared of 0.62. The amplitudes for the IG group unlike the EG and CTRL group also decrease over intervention. There frequencies also drift further away from the ideal frequency post intervention.

Figure 7 shows the distribution of all individuals averaged on all foot tapping trials. The FFT of an ideal foot tapping trial with equal amplitude in the slow and fast beats as shown in Figure 1D & E. From the post results, the amplitude between the fast beat (1Hz) to the slow beat (2Hz) is roughly 2:5. This ratio arises from the slow tap amplitude containing all amplitude from all 5 taps, and the fast tap amplitude just from the two faster taps. Further along this line from the origin represents a higher degree deflection during foot tapping and moving below or above the line represents higher amplitudes either during the slow or the fast tapping parts of the rhythm respectively. For the frequencies, the trials closer to the 1 Hz for slow taps and 2Hz for fast taps, observed the correct timing during the trial. The mean cluster center of each dance group is shown in diamond in Figure 7. Over the intervention, the two dance groups amplitudes approached the 2:5 ratio, where the r-squared to the line y=2/5x increased from pre to post from 0.17 to 0.61 in the IG group and 0.04 to 0.59 in the EG group and the control group r-squared decreased from 0.27 to 0.03 for the ET task. This finding suggests that the dance groups were more coordinated post intervention on the foot tapping dance task and were able to have equal amplitude across TANGO beat. However, the IG group, unlike the EG and control group also showed a shift towards the origin suggesting a smaller foot deflections post intervention. The IG group frequency also shifted further from the ideal beat frequency post intervention.

## 4. DISCUSSION

This study was designed to determine whether IG or EG training results in differential engagement of pathways in people with mild to moderate PD who are known to have impaired STC pathways and relatively preserved CTC pathways. Given the training, adapted tango, consistently improve clinical mobility outcomes (Hackney et al, 2007; Hackney and Earhart 2009; Hackney and Earhart 2010; McKee and Hackney 2013; McKay et al., 2016), we sought to determine the neural changes that patients use if they exclusively train in EG or IG training. A major question in rehabilitation is whether effective therapies (like Adapted tango that show clinical benefit) lead to restoration within impaired circuits (e.g., the STC circuit in this study), in other words-restoring neurological function, or whether alternative compensatory circuits are engaged more after successful training. The EG group had the largest changes in all behavioral outcomes on PD, motor, cognitive, and psychosocial variables. The IG group also had improved in motor and PD variables. Moreover, as expected, both dance group participants exceeded the minimal clinical important difference (MCID) (3 points) for the MDS-UPDRS-III score (Hauser et al., 2011; Horvath et al., 2015). This study showed initial evidence of neural pathways differentially affected by IG versus EG training. The neural changes indicated that only the EG group had significant increase in recruitment of the CTC pathway and increased activation in the motor cortex. The IG group showed the least changes in neural measures in all pathways, compared to other groups. The control group had a significant increase in the recruitment of striatal pathways compared to the two dance groups, and also showed less clinically measured depression, but no changes in other regions of the brain.

### 4.1 Activation during Baseline Task Activation

The regions that were activated during baseline confirm published results and our understanding of motor circuitry. The motor cortex is topologically organized, and the primary area that was activated in the left motor cortex is linked to the foot region and corresponds to the contralateral right foot tapping task (Trinastic et al., 2010; Marieb and Hoehn 2007; Ciccarelli et al., 2005; Goldman-Rakic et al., 1992; Drucker et al., 2019). Moreover, the areas in the dlPFC and motor planning areas match earlier characterizations of the motor circuit (Goldman-Rakic et al., 1992; Halsband et al., 1994; Trinastic et al., 2010). The activation in the putamen is also present as expected from previous studies, confirming that these striatal areas are more active during task in PD due to compensation (Ceresa et al., 2006). The areas that were active during this foot tapping task in the cerebellum lobules VIIIa/b are also active during lower limb tasks (Ciccarelli et al., 2006). Moreover, the two tasks displayed selectivity in activating the ET vs IT circuitry as shown in Drucker et al., 2018, where the IT task shows more significant recruitment in the putamen, and the ET task is faster in activating in the cerebellum. Therefore, the results present a circuit that has been well characterized in the motor literature, where the parietal lobe functions as sensory integration, the dlPFC functions as the executive command, and the STC and CTC circuits in tandem with M1 functionally coordinate the movement (Goldman-Rakic et al., 1992; Ciccarelli et al., 2005; Halsband et al., 1994; Marieb and Hoehn 2007, Trinastic et al., 2010).

### 4.2 Changes due to Striatal Motor Neural Rehabilitation

We hypothesize that the leaders would show more STC activation post intervention, because leading in partnered dance requires planning, updating and goal selection, all executive functions to determine step direction, timing and amplitude, which they then would self-initiate (Ceresa et al., 2006; Ciccarelli et al., 2005; Lewis et al., 2008). The leader group, unlike the controls, showed significant clinical improvement in UPDRS III and motor measures such as the balance and the distance they are able to walk during a six-minute interval, indicating that the IG dance training had positive benefits to the PD participants. These results complement older studies that have shown positive behavioral benefits of using striatal focused rehabilitation (Dibble et al., 2004). However, the fMRI analysis revealed that the IG group showed the least amount of changes in activation during the in-scanner fMRI task performance. Surprisingly, the STC circuity was most active post intervention the control group. From an earlier study (Palmer et al., 2009), the striatal areas are more active in PD participants than healthy controls for the same task. Palmer et al., 2009 suggested this finding may be due to a compensatory mechanism. Therefore, having a smaller response during the task might mean that the dance group gained a larger range of response by recruiting other circuits, whereas the control group might have utilized the striatum and therefore had a larger response during task. This possibility could also explain why changes are not only in the left putamen that was more active during baseline task performance but also in the contralateral right putamen. Behaviorally the control group had no changes in any of the behavioral variables suggesting that these changes had no relevance in improving motor outcomes. The IG group may have performed worse during in scanner foot tapping task compared to the other two groups, but they had improvements in behavioral tasks performed outside of the scanner. Therefore, from the task fMRI experiment we conclude that by directly training the striatal pathway, no improvement was seen in the overall activation during gait initiation in the STC or primary motor cortex pathways and might even contribute to a decay in performance of the in-scanner foot tapping task. These results must be interpreted cautiously however since seem to support the futility of improving the STC circuit through rehabilitative means. The changes in other behavioral outcomes such as postural control, balance, and the overall UPDRS score were probably due to changes in pathways other than the STC circuits.

### 4.3 Changes due to Cerebellar Motor Neural Rehabilitation

The EG group showed the most signs of improvement using the behavioral and clinical measures. Not only did they improve in motor variables such as UPDRS III or Freezing of Gait, but they also showed improvement in working memory tasks (Brooks Spatial Memory and Corsi Product), and significant reductions in depression according to the Beck’s depression score. This is not surprising as sensory cuing has been shown to improve motor variables such as gait speed and later studies showed benefits of multimodal rehabilitation therapy (Hackney et al., 2015, Dibble et al., 2004). The leader group also improved in the motor variables which might be due to a similar mechanism. The fMRI analysis also showed that only the follower group had a significant increase of activity in cerebellar circuits, at both the input and the output of the circuit (the pons and DCN regions respectively Figure 5). This observation is consistent with our hypothesis that EG movement training, recruits the Cerebellar structures, especially near the VIIIA/B region previously identified as part of the sensory motor circuit (Ciccarelli et al., 2006). Moreover, these changes in the cerebellum translated into significant changes in other parts of the motor circuit, including motor planning regions and the primary motor cortex M1. This observation is contrary to the IG or the control groups that showed no significant changes in the motor areas. Since these changes were only observed using the atlas approach and not the voxel approach, the EG training might have improved activity over a widespread region of the motor cortex, unlike changes in the subcortical areas where the changes were relatively localized. The EG group also showed the most improvement during the foot tapping task, further suggesting that the cerebellar changes led to changes in the primary motor cortex and then manifest in the behavioral responses.

### 4.4 Limitations

The EG and IG training roles served as imperfect proxies for IG and EG movements, and there may be no pure form of EG or IG training. The results from this study must be interpreted cautiously. Small, and unequal sample sizes may have reduced power to detect effects. Larger samples are necessary to produce more definitive results. The in-scanner foot tapping task (ET and IT) showed very few behavioral differences between the individuals. Similar to working memory tasks using more N-backs, making the foot tapping task more difficult, i.e more complicated rhythm, could improve the study as well. The results from this inquiry must be interpreted cautiously since our sample size is not very large.

## 5. CONCLUSION

This study is one of the first attempts to dissociate EG from IG training and examine its relative effects on related or associated pathways. Our study showed no enhanced activity in the STC pathway following IG training in our defined PD groups. Robust results in the CTC pathway, and associated areas in the EG group indicate possibly greater potential in neural plasticity after EG training. Benefits from motor rehabilitation are more nuanced, and likely rely heavily on the underlying circuitry for improvement. When planning treatments, clinicians should be cognizant of the neural pathways they are targeting and have a mechanistic hypothesis on how their intervention would improve health outcomes. Improved motor benefits in PD may result from highly selective rehabilitation movements.

## Supporting information

clinical trail id

## Data Availability

The authors confirm that, for approved reasons, some access restrictions apply to the data underlying the findings. Public data deposition is not ethical or legal and would compromise patient privacy. Data are, in part, housed in the VINCI Veterans Affairs (VA) Informatics and Computing Infrastructure (VINCI), a secure, virtual computing environment developed through a partnership between the VA Office of Information Technology (OI&T) and the Veterans Health Administration's Office of Research and Development (VHA ORD). Data are also stored on a VA secured server under a Data Usage Agreement with Emory University. VINCI is a partner with the Corporate Data Warehouse (CDW) and hosts all data available through CDW. VHA National Data Services (NDS) authorizes research access to patient data. Data are only available for researchers who meet the criteria for access to confidential data. Contact.

## Acknowledgements

We thank the volunteers and participants for their time and effort devoted to this study.

## Author Contributions

1. Research Project: A. Conception: AK, MEH, KS, BC, DC, SW, JD, ME Organization, MEH; KM, VK, GK, C. Execution: AK, MEH, JD, KM, AB, ME
2. Statistical Analysis: A. Design, AK, MEH, JD, VK, KM, LK B. Execution, AK, JD, MEH C. Review and Critique: KS, BC, VK, SW, DC, GK, ME
3. Manuscript: A. Writing of the final draft, AK, MEH B. Review and Critique: KS, BC, SW, DC, ME, GK, VK

