## supplementary table for "Neural Correlates of Effects of Internally versus Externally Guided Partnered Rehabilitative Tango for People with Parkinson’s Disease"

### Supplementary Tables:

#### ROI Atlas Significant table

| Test | ROI # | Brainetome area | beta | grey matter volume |
|---|---|---|---|---|
| EG pre vs post IT | 8 | 'superior frontal gyrus, part 4 (dorsolateral area 6), right', | 0.773 | 17388 |
| | 11 | 'superior frontal gyrus, part 6 (medial area 9), left', | 0.793 | 18630 |
| | 12 | 'superior frontal gyrus, part 6 (medial area 9), right', | 0.806 | 24543 |
| | 17 | 'middle frontal gyrus, part 2 (inferior frontal junction), left', | 0.819 | 22140 |
| | 35 | 'inferior frontal gyrus, part 4 (rostral area 45), left', | 1.3 | 10665 |
| | 67 | 'paracentral lobule, part 2 (area 4 lower limb), left', | 0.86 | 13662 |
| EG pre vs post ET | 20 | 'middle frontal gyrus, part 3 (area 46), right', | 0.934 | 28188 |
| | 21 | 'middle frontal gyrus, part 4 (ventral area 9/46 ), left' | 0.812 | 27864 |
| | 31 | 'inferior frontal gyrus, part 2 (inferior frontal sulcus), left', | 0.864 | 11124 |
| | 35 | 'inferior frontal gyrus, part 4 (rostral area 45), left', | 1 | 10665 |
| | 50 | 'orbital gyrus, part 5 (area 13), right', | -1.7 | 22626 |
| | 67 | 'paracentral lobule, part 2 (area 4 lower limb), left', | 1.1 | 13662 |
| | 180 | 'cingulate gyrus, part 3 (pregenual area 32), right', | 0.9 | 10665 |
| | 261 | 'Cerebellar lobule VIIb, vermis', | -1.5 | 729 |
| IG pre vs post IT | 16 | middle frontal gyrus, part 1 (dorsal area 9/46), right', | -0.8 | 27567 |
| | 24 | 'middle frontal gyrus, part 5 (ventrolateral area 8), right', | -0.8 | 22437 |
| | 40 | 'inferior frontal gyrus, part 6 (ventral area 44), right', | -0.9 | 8262 |
| | 54 | 'precentral gyrus, part 1 (area 4 head and face), right', | -0.9 | 14175 |
| | 64 | 'precentral gyrus, part 2 (caudal dorsolateral area 6), left', | -1.3 | 18819 |
| | 81 | 'middle temporal gyrus, part 1 (caudal area 21), left', | -1.4 | 14472 |
| | 99 | 'inferior temporal gyrus, part 6 (caudolateral of area 20), left', | -1.6 | 14310 |
| | 100 | 'inferior temporal gyrus, part 6 (caudolateral of area 20), right', | -1.6 | 10962 |
| | 126 | 'superior parietal lobule, part 1 (rostral area 7), right', | -1.2 | 14904 |
| | 128 | 'superior parietal lobule, part 2 (caudal area 7), right', | -1.5 | 14094 |
| | 134 | 'superior parietal lobule, part 5 (intraparietal area 7), right', | -1.4 | 11853 |
| | 136 | 'inferior parietal lobule, part 1 (caudal area 39), right', | -1.3 | 33021 |
| | 144 | 'inferior parietal lobule, part 5 (rostroventral area 39), right', | -1 | 37098 |
| | 191 | 'cuneus, part 2 (rostral cuneus gyrus), left', | -1.2 | 23706 |
| | 192 | 'cuneus, part 2 (rostral cuneus gyrus), right', | -1.1 | 22248 |
| | 200 | 'occipital gyrus, part 1 (middle occipital gyrus), right', | -0.95 | 23220 |
| | 210 | 'superior occipital gyrus, part 2 (lateral superior occipital gyrus), right', | -1 | 20979 |
| IG pre vs post ET | 1 | 'superior frontal gyrus, part 1 (medial area 8), left', | -1 | 21033 |
| | 2 | 'superior frontal gyrus, part 1 (medial area 8), right', | -0.8 | 23706 |
| | 4 | 'superior frontal gyrus, part 2 (dorsolateral area 8), right', | -0.7 | 20007 |
| | 10 | 'superior frontal gyrus, part 5 (medial area 6), right', | -0.8 | 21276 |

| | | | |
|---|---|---|---|
| 16 | middle frontal gyrus, part 1 (dorsal area 9/46), right', | -0.7 | 27567 |
| 24 | middle frontal gyrus, part 5 (ventrolateral area 8), right', | -0.7 | 22437 |
| 54 | precentral gyrus, part 1 (area 4 head and face), right', | -0.8 | 14175 |
| 56 | 'precentral gyrus, part 2 (caudal dorsolateral area 6), right', | -0.6 | 23328 |
| 64 | 'precentral gyrus, part 6 (caudal ventrolateral area 6), right', | -1 | 18819 |
| 65 | 'paracentral lobule, part 1 (area1/2/3 lower limb), left', | -1 | 9720 |
| 66 | 'paracentral lobule, part 1 (area1/2/3 lower limb), right', | -0.8 | 12474 |
| 68 | paracentral lobule, part 2 (area 4 lower limb), right', | -1 | 16200 |
| 81 | middle temporal gyrus, part 1 (caudal area 21), left', | -2.1 | 14472 |
| 93 | 'inferior temporal gyrus, part 3 (rostral area 20), left', | -2.4 | 12987 |
| 99 | inferior temporal gyrus, part 6 (caudolateral of area 20), left', | -2.1 | 14310 |
| 100 | inferior temporal gyrus, part 6 (caudolateral of area 20), right', | -1.9 | 10962 |
| 106 | 'fusiform gyrus, part 2 (medioventral area 37), right', | -1.4 | 19980 |
| 126 | superior parietal lobule, part 1 (rostral area 7), right', | -1.3 | 14904 |
| 128 | superior parietal lobule, part 2 (caudal area 7), right', | -1.6 | 14094 |
| 130 | 'superior parietal lobule, part 3 (lateral area 5), right', | -0.9 | 10908 |
| 133 | 'superior parietal lobule, part 5 (intraparietal area 7), left', | -1 | 11475 |
| 134 | superior parietal lobule, part 5 (intraparietal area 7), right', | -1.5 | 11853 |
| 136 | inferior parietal lobule, part 1 (caudal area 39), right', | -1.6 | 33021 |
| 138 | 'inferior parietal lobule, part 2 (rostrodorsal area 39), right', | -1.4 | 28458 |
| 148 | 'precuneus, part 1 (medial area 7), right', | -1.5 | 12447 |
| 150 | 'precuneus, part 2 (medial area 5), right', | -1.2 | 18333 |
| 161 | 'postcentral gyrus, part 4 (area1/2/3 trunk), left', | -1 | 19413 |
| 189 | cuneus, part 1 (caudal lingual gyrus), left', | -1.8 | 13554 |
| 191 | cuneus, part 2 (rostral cuneus gyrus), left', | -1.3 | 23706 |
| 192 | cuneus, part 2 (rostral cuneus gyrus), right', | -1.4 | 22248 |
| 193 | 'cuneus, part 3 (caudal cuneus gyrus), left', | -2.3 | 17496 |
| 194 | 'cuneus, part 3 (caudal cuneus gyrus), right', | -1.3 | 15471 |
| 197 | cuneus, part 5 (ventomedial parietooccipital sulcus), left', | -1.1 | 27351 |
| 200 | occipital gyrus, part 1 (middle occipital gyrus), right', | -1.6 | 23220 |
| 203 | 'occipital gyrus, part 3 (occipital polar cortex), left', | -2 | 28134 |
| 204 | 'occipital gyrus, part 3 (occipital polar cortex), right', | -2.1 | 28809 |
| 206 | 'occipital gyrus, part 4 (inferior occipital gyrus), right', | -2.4 | 26325 |
| 210 | superior occipital gyrus, part 2 (lateral superior occipital gyrus), right', | -1.3 | 20979 |
| 257 | 'Cerebellar Crus II, left', | -1.8 | 56511 |
| 259 | 'Cerebellar Crus II, right', | -1.9 | 50571 |
| 260 | 'Cerebellar lobule VIIb, left', | -2.1 | 23166 |
| 262 | Cerebellar lobule VIIb, right', | -2.3 | 24840 |
| 263 | 'Cerebellar lobule VIIIa, left', | -2.1 | 23274 |
| 264 | 'Cerebellar lobule VIIIa, vermis', | -1.7 | 6075 |
| 265 | 'Cerebellar lobule VIIIa, right', | -2.2 | 23598 |
| 267 | 'Cerebellar lobule VIIIb, vermis', | -1.9 | 3645 |
| 268 | 'Cerebellar lobule VIIIb, right', | -1.9 | 19305 |
| 269 | 'Cerebellar lobule IX, left', | -1.5 | 14769 |

| | | | | |
|---|---|---|---|---|
| ctrl pre vs post IT | 23 | 'middle frontal gyrus, part 5 (ventrolateral area 8), left', | 0.6 | 27486 |
| | 81 | middle temporal gyrus, part 1 (caudal area 21), left', | 1.7 | 14472 |
| | 82 | 'middle temporal gyrus, part 1 (caudal area 21), right', | 1.1 | 19278 |
| | 89 | 'inferior temporal gyrus, part 1 (intermediate ventral area 20), left', | 1.6 | 8316 |
| | 95 | 'inferior temporal gyrus, part 4 (intermediate lateral area 20), left', | 1.3 | 12744 |
| | 99 | inferior temporal gyrus, part 6 (caudolateral of area 20), left', | 1.6 | 14310 |
| | 108 | 'fusiform gyrus, part 3 (ventrolatral area 37), right', | 1.3 | 24300 |
| | 203 | 'occipital gyrus, part 3 (occipital polar cortex), left', | 1.1 | 28134 |
| | 263 | 'Cerebellar lobule VIIIa, left', | 1.3 | 23274 |
| CTRL pre vs post ET | 134 | superior parietal lobule, part 5 (intraparietal area 7), right', | 1.1 | 11853 |
| | 205 | 'occipital gyrus, part 4 (inferior occipital gyrus), left', | 1.5 | 29079 |
| EG vs CTRL IT | | No changes | | |
| EG vs CTRL ET | 67 | 'paracentral lobule, part 2 (area 4 lower limb), left', | 0.976 | 13662 |
| IG vs CTRL IT | 1 | 'superior frontal gyrus, part 1 (medial area 8), left', | -0.8 | 21033 |
| | 64 | 'precentral gyrus, part 6 (caudal ventrolateral area 6), right', | -1 | 18819 |
| | 81 | middle temporal gyrus, part 1 (caudal area 21), left', | -1.9 | 14472 |
| | 99 | inferior temporal gyrus, part 6 (caudolateral of area 20), left', | -1.9 | 14310 |
| | 134 | superior parietal lobule, part 5 (intraparietal area 7), right', | -1 | 11853 |
| IG vs CTRL ET | 128 | superior parietal lobule, part 2 (caudal area 7), right', | -1.5 | 14094 |
| | 134 | superior parietal lobule, part 5 (intraparietal area 7), right', | -1.6 | 11853 |
| | 148 | 'precuneus, part 1 (medial area 7), right', | -1.3 | 12447 |
| | 181 | 'cingulate gyrus, part 4 (ventral area 23), left', | -1.2 | 9612 |
| | 189 | cuneus, part 1 (caudal lingual gyrus), left', | -1.7 | 13554 |
| | 192 | cuneus, part 2 (rostral cuneus gyrus), right', | -1.2 | 22248 |
| | 193 | 'cuneus, part 3 (caudal cuneus gyrus), left', | -1.6 | 17496 |
| | 200 | occipital gyrus, part 1 (middle occipital gyrus), right', | -1.1 | 23220 |
| | 203 | 'occipital gyrus, part 3 (occipital polar cortex), left', | -1.3 | 28134 |
| | 205 | 'occipital gyrus, part 4 (inferior occipital gyrus), left', | -1.9 | 29079 |
| | 206 | 'occipital gyrus, part 4 (inferior occipital gyrus), right', | -1.6 | 26325 |
| EG vs IG IT | 2 | 'superior frontal gyrus, part 1 (medial area 8), right', | 0.8 | 23706 |
| | 8 | superior frontal gyrus, part 4 (dorsolateral area 6), right', | 0.7 | 17388 |
| | 11 | 'superior frontal gyrus, part 6 (medial area 9), left', | 0.8 | 18630 |
| | 12 | superior frontal gyrus, part 6 (medial area 9), right', | 0.8 | 24543 |
| | 16 | middle frontal gyrus, part 1 (dorsal area 9/46), right', | 0.8 | 27567 |
| | 26 | 'middle frontal gyrus, part 6 (ventrolateral area 6), right', | 0.8 | 17820 |
| | 37 | 'inferior frontal gyrus, part 5 (opercular area 44), left', | 0.7 | 12042 |
| | 40 | inferior frontal gyrus, part 6 (ventral area 44), right', | 0.9 | 8262 |
| | 56 | 'precentral gyrus, part 2 (caudal dorsolateral area 6), right', | 0.7 | 23328 |

| | | | | |
|---|---|---|---|---|
| | 64 | 'precentral gyrus, part 6 (caudal ventrolateral area 6), right', | 1.2 | 18819 |
| EG vs IG ET | 2 | superior frontal gyrus, part 1 (medial area 8), right', | 0.7 | 23706 |
| | 4 | 'superior frontal gyrus, part 2 (dorsolateral area 8), right', | 0.7 | 20007 |
| | 10 | 'superior frontal gyrus, part 5 (medial area 6), right', | 0.8 | 21276 |
| | 57 | 'precentral gyrus, part 3 (area 4 upper limb), left', | 0.8 | 16308 |
| | 59 | 'precentral gyrus, part 4 (area 4 trunk), left', | 0.9 | 7425 |
| | 64 | 'precentral gyrus, part 6 (caudal ventrolateral area 6), right', | 0.8 | 18819 |
| | 65 | 'paracentral lobule, part 1 (area1/2/3 lower limb), left', | 1.1 | 9720 |
| | 66 | 'paracentral lobule, part 1 (area1/2/3 lower limb), right', | 0.9 | 12474 |
| | 67 | 'paracentral lobule, part 2 (area 4 lower limb), left', | 1.2 | 13662 |
| | 68 | paracentral lobule, part 2 (area 4 lower limb), right', | 1.1 | 16200 |
| | 71 | 'superior temporal gyrus, part 2 (area 41/42), left', | 0.9 | 12636 |
| | 148 | 'precuneus, part 1 (medial area 7), right', | 1.5 | 12447 |
| | 153 | 'precuneus, part 4 (area 31), left', | 0.9 | 21249 |
| | 154 | 'precuneus, part 4 (area 31), right', | 0.9 | 26757 |
| | 161 | 'postcentral gyrus, part 4 (area1/2/3 trunk), left', | 0.9 | 19413 |
| | 193 | 'cuneus, part 3 (caudal cuneus gyrus), left', | 1.7 | 17496 |
| | 265 | 'Cerebellar lobule VIIIa, right', | 2.3 | 23598 |
