## Supplementary material for "Neural Correlates of Effects of Internally versus Externally Guided Partnered Rehabilitative Tango for People with Parkinson’s Disease": clinical trail id

**ClinicalTrials.gov Protocol Registration and Results System (PRS) Receipt**
Release Date: June 28, 2021

**ClinicalTrials.gov ID: NCT02457832**

### Study Identification

Unique Protocol ID: N0870-W

Brief Title: Motor Training in PD

Official Title: Optimizing Motor Training in Parkinson Disease Through Neural Mechanisms (NEURODEGEN)

Secondary IDs:

### Study Status

Record Verification: June 2021

Overall Status: Active, not recruiting

Study Start: November 3, 2014 [Actual]

Primary Completion: December 31, 2022 [Anticipated]

Study Completion: December 31, 2023 [Anticipated]

### Sponsor/Collaborators

Sponsor: VA Office of Research and Development

Responsible Party: Sponsor

Collaborators:

### Oversight

U.S. FDA-regulated Drug: No

U.S. FDA-regulated Device: No

Unapproved/Uncleared Device: No

U.S. FDA IND/IDE: No

Human Subjects Review: Board Status: Approved
Approval Number: 00060613
Board Name: Emory University Institutional Review Board
Board Affiliation: Emory University

Address:

Atlanta VA Medical and Rehab Center, Decatur
1670 Clairmont Road

Decatur, GA 30033
Country: USA

Data Monitoring: No

### Study Description

Brief Summary: The purpose of this research study is to learn more about brain activity when individuals with and without Parkinson disease (PD) move their lower limbs. The investigators also want to see if and how two different types of partnered dance affect brain activity in individuals with and without PD. Testing will take place at the Atlanta VA Medical Center and at Emory University. The investigators expect to enroll about 140 people for this study over a five-year period.

Detailed Description: Persons with Parkinson's disease (PD) have impaired mobility, which adversely affects their quality of life. The effectiveness of adapted tango dance, in which participants both lead (internally guide: IG) and follow (externally guide: EG) movement has been shown. To improve outcomes in those with PD, the underlying brain mechanisms for both motor impairments and improvement must be studied. IG and EG movements have distinct brain activity patterns. Individuals with PD have trouble with IG movement but this problem is helped by strategies used while "leading." During "following", participants with PD can use many external cues, which helps movement in PD, because EG tasks bypass the basal ganglia, the part of the brain affected by PD. In older persons with PD, the investigators aim to:

- determine brain activation patterns during IG and EG foot movement.
- look into effects of IG and EG training on brain activation along with mobility improvements.

The investigators will begin with a functional Magnetic Resonance Imaging test in a scanner. The investigators will look at brain area correlates of a clinically-used foot-tapping task, during IG and EG conditions in older persons with and without PD. Then, the investigators will assess the relative effectiveness of IG versus EG training during an adapted tango class, compared to a group that participates in health education, for improved mobility and foot tapping. Participants with PD will be assessed for disease severity. They will receive tests of outcome measures while "OFF" and "ON" PD-specific medications at the following time points:

- 1 week before training
- 1 week after training
- 1 month after training Participants must attend 20 lessons of IG or EG adapted tango in 12 weeks, taught by an experienced instructor. In the functional MRI (fMRI) scanner, the investigators will assess participants for improved foot tapping after training. The investigators will also look at changes in activation in specific brain circuits along with training effects upon mobility.

The long-term goal is to improve motor training as much as possible for persons with PD by understanding foot movement brain circuitry in PD as well as brain changes in circuitry through which training is effective. This work proposes to illumine information about brain function that is very important to continued progress in rehabilitative care of persons with PD.

### Conditions

Conditions: Parkinson Disease

| | |
|---|---|
| Keywords: | Magnetic Resonance Imaging<br>Neurology<br>Neurosciences<br>Physical Medicine<br>Parkinson Disease<br>Rehabilitation |

### Study Design

| | |
|---|---|
| Study Type: | Interventional |
| Primary Purpose: | Supportive Care |
| Study Phase: | N/A |
| Interventional Study Model: | Parallel Assignment |
| Number of Arms: | 4 |
| Masking: | None (Open Label) |
| Allocation: | Randomized |
| Enrollment: | 99 [Anticipated] |

### Arms and Interventions

| Arms | Assigned Interventions |
|---|---|
| Experimental: Internal guidance training (IG)<br>Adapted tango dancing is a sophisticated, yet accessible system of tactile communication that conveys motor intentions and goals between a leader and follower. Those in IG training will choose direction, timing and amplitude of each successive step, and will communicate this information to their partner through moving their frame and center of mass. | Behavioral: Adapted Tango Dancing<br>Composed of simple steps, tango involves frequent movement initiation and cessation, multi-directional perturbations and varied rhythms. Participants focus on trunk control and stepping strategies, coordination, somatosensory awareness, attention to partner, path of movement, and aesthetics. Sessions will begin with a typical dance class warm-up consisting of breathing, limbering and postural alignment to upbeat music. Novel step elements will be introduced every class period. Those with PD will partner with an individual without PD. After novel step introduction, the instructor will present rhythmic training, which is indispensable to partnered dancing. Participants will learn 'typical' rhythms from tango and Latin dances, based upon the system of quicks (Q) and slows (S), ubiquitously used in ballroom dance training to understand the temporal relationship of movement to music. |
| Experimental: Externally guided training (EG)<br>Those in EG will learn to attend to sensory cues for movement direction, timing and amplitude of steps, communicated from their partner to them via the frame and center of mass. The 'follower' will wait to receive the movement cue before moving. | Behavioral: Adapted Tango Dancing<br>Composed of simple steps, tango involves frequent movement initiation and cessation, multi-directional perturbations and varied rhythms. Participants focus on trunk control and stepping strategies, coordination, somatosensory awareness, attention to partner, path of movement, and aesthetics. Sessions will begin with a typical dance class warm-up consisting of breathing, limbering and postural alignment to upbeat music. Novel step elements will be introduced every class period. Those with PD will partner with an individual without PD. After novel step introduction, the instructor will present rhythmic training, which is indispensable to partnered dancing. Participants will learn 'typical' |

| Arms | Assigned Interventions |
|---|---|
| | rhythms from tango and Latin dances, based upon the system of quicks (Q) and slows (S), ubiquitously used in ballroom dance training to understand the temporal relationship of movement to music. |
| Active Comparator: Behavioral Control (BC)<br>BC participants will attend group health education sessions adapted to the needs of individuals with PD, about pharmacological management, nutrition, sleep disorders, cognitive deficits, bereavement coping, mobility, balance and home safety. Participants in this training will be instructed not to change their habitual exercise routines. After completing health education, participants will be assigned to an IG or EG training class but will not undergo evaluations. | Behavioral: Behavioral Control (BC) Condition<br>Group health education sessions adapted to the needs of individuals with PD, about pharmacological management, nutrition, sleep disorders, cognitive deficits, bereavement coping, mobility, balance and home safety. |
| No Intervention: Normal Control (NC)<br>Age-matched controls without Parkinson's disease will come in for a single assessment including MRI. | |

### Outcome Measures

Primary Outcome Measure:

1. Percent signal change
   For the IG and EG tasks for the MRI, the investigators want to determine and distinguish circuits involved in IG and EG foot-tapping networks in participants with and without PD.

   [Time Frame: 12 weeks]
2. Connectivity strength
   Changes in average connectivity strength across striatal-thalamo-cortical (STC) and cerebello-thalamo-cortical (CTC) circuits, as measured by average cross correlation coefficient between the seed regions of the circuits.

   [Time Frame: 12 weeks]

### Eligibility

Minimum Age: 40 Years

Maximum Age:

Sex: All

Gender Based: No

Accepts Healthy Volunteers: Yes

Criteria: Inclusion Criteria:

- Age 40 - 70 years
- Willingness to spend 1-h in a scanner
- Able to walk with or without an assistive device 10 feet
- Best corrected/aided acuity better than 20/70 in the better eye
- Absence of dementia or vascular cognitive impairment
- Absence of primary memory deficits

Exclusion Criteria:

- Deep brain stimulator implants, Metallic implants, fragments, or pacemakers
- Montreal Cognitive Assessment (MocA) score < 24
- Pure-tone threshold sensitivity > 40 dB

- Peripheral neuropathy
- Untreated Major Depression
- History of stroke, or traumatic brain injury

### Contacts/Locations

Central Contact Person: Madeleine E Hackney, PhD


Central Contact Backup: Laura Britan Lang, MPH


Study Officials: Madeleine E. Hackney, PhD
Study Principal Investigator
Atlanta VA Medical and Rehab Center, Decatur, GA

Locations: **United States, Georgia**
Atlanta VA Medical and Rehab Center, Decatur, GA
Decatur, Georgia, United States, 30033
Contact: Ashley N Scales 404-321-6111 Ext. 23952
Contact: Laura Britan Lang, MPH (404) 321-6111 Ext. 7027

Principal Investigator: Madeleine E. Hackney, PhD

### IPDSharing

Plan to Share IPD: No

### References

Citations:

Links:

Available IPD/Information:
